# Exploring Risks of Human Challenge Trials for COVID-19

**DOI:** 10.1101/2020.11.19.20234658

**Authors:** David Manheim, Witold Więcek, Virginia Schmit, Josh Morrison, 1Day Sooner Research Team

## Abstract

Human Challenge Trials (HCTs) are a potential method to accelerate development of vaccines and therapeutics. However, HCTs for COVID-19 pose ethical and practical challenges, in part due to the unclear and developing risks. In this paper, we introduce an interactive model for exploring some risks of a SARS-COV-2 dosing study, a prerequisite for any COVID-19 challenge trials. The risk estimates we use are based on a Bayesian evidence synthesis model which can incorporate new data on infection fatality risks (IFRs) to patients, and infer rates of hospitalization. The model estimates individual risk, which we then extrapolate to overall mortality and hospitalization risk in a dosing study. We provide a web tool to explore risk under different study designs.

Based on the Bayesian model, IFR for someone between 20 and 30 years of age is 15.1 in 100,000, with a 95% uncertainty interval from 11.8 to 19.2, while risk of hospitalization is 130 per 100,000 (100 to 160). However, risk will be reduced in an HCT via screening for comorbidities, selecting lower-risk population, and providing treatment. Accounting for this with stronger assumptions, we project the fatality risk to be as low as 2.5 per 100,000 (1.6 to 3.9) and the hospitalization risk to be 22.0 per 100,000 (14.0 to 33.7). We therefore find a 50-person dosing trial has a 99.74% (99.8% to 99.9%) chance of no fatalities, and a 98.9% (98.3% to 99.3%) probability of no cases requiring hospitalization.

## 1 INTRODUCTION

As of February 1, 2021 severe acute respiratory syndrome coronavirus 2 (SARS-CoV-2) has led to over one hundred million confirmed infections worldwide, and over 2 million deaths. Despite the recent approval of vaccines, McKinsey estimates that global vaccine production will reach eight billion doses in 2021 – and because the vaccines require two doses, this is only enough to vaccinate a bit over half of the global population (Agrawal, Conway, Heller, Sabow, & Tolub, 2020). However, now that vaccines are approved, testing of additional vaccines with large-scale field trials is difficult and morally fraught. Not only that, but a variety of critical questions are emerging about the extant vaccines, including the effectiveness of single doses, and the effectiveness against novel strains of COVID.

Rapid answers have been, and will continue to be, critical in the management of disease burden and mitigating impacts of COVID-19. Different paths to producing relevant clinical data exist, but field trials are slow and require large subject populations to discern therapeutic effects (Jamrozik & Selgelid, 2020b), while observational studies of effectiveness are similarly slow, and not randomized. In order to accelerate testing of vaccines, several authors and institutions have proposed intentionally exposing human subjects to SARS-CoV-2 to test novel interventions; to this date, nearly forty thousands of people have expressed interest in volunteering for this task (1Day Sooner, 2020). Such human challenge trials (HCTs) have been useful in the past (Jamrozik & Selgelid, 2021) to develop vaccines and treatments for other infectious diseases such as malaria, cholera, respiratory syncytial virus (RSV), (Roestenberg, Hoogerwerf, Ferreira, Mordmüller, & Yazdanbakhsh, 2018; Gómez-Pérez et al., 2015) influenza, (Treanor et al., 1999) and dengue fever (Larsen, Whitehead, & Durbin, 2015). While the changing situation has led some to question the need for HCTs, others have defended their usefulness (Turk, 2021). The usefulness is because HCTs can be used to answer a variety of questions (Nguyen et al., 2020), including many not addressed by the availability of current vaccines (Steuwer, Jamrozik, & Eyal, 2021; Ducarmon, Kuijper, & Olle, 2021). However, two ethical problems have been raised which stand in the way of conducting HCTs for SARS-CoV-2 infection. First, given lack of rescue therapies and our limited understanding of COVID-19’s risks, it is difficult to weigh the likely impacts of these studies on volunteers against the benefit to society, or to obtain informed consent from volunteers (Palmer & Schuck, 2020). Second, we do not know what viral dose of SARS-CoV-2 should be given to volunteers.

To help address both concerns, we developed a model to help assess the risks participants will face in a hypothetical dosing study for COVID-19, one similar to hVIVO’s soon-to-be-launched trial (Turk, 2021). This model uses data from a non-systematic review of data on COVID-19 risks (mortality and infection rates) and describes risks for individuals as well as the overall study risk. As both clarification on viral dose and infection risks are essential before starting HCTs, this work can help inform policymakers and potential volunteers about some risks concerning the process of using HCTs to accelerate vaccine and therapeutic development.

## 2 METHODOLOGY

We developed a three-component tool to understand and explain the relevant risks. The first component quantifies risk of COVID-19 mortality by using a Bayesian evidence synthesis model; the second uses that estimate, along with other data on gender-specific mortality and hospitalization risks to simulate the risk of a study with given characteristics; the third is a front end tool for allowing interactive exploration of risks from a study or to an individual.

### 2.1 Bayesian evidence synthesis model

We use a Bayesian meta-analysis approach to obtain an estimate of mortality risk. This form of modelling combines different sources of evidence to characterise both mean and dispersion in a given statistic of interest. In our case, we use age-specific, location-specific death and prevalence data to generate an estimate of the infection fatality risk (IFR), i.e. the probability that a person infected with COVID-19 will die, accounting for age; we also provide an estimate of hospitalization risk. These two estimates allow us to understand risk reduction in individuals who would participate in an HCT.

We use Bayesian methods because they allow us to best account for heterogeneity in IFRs across age groups, different countries and regions. Characterising this heterogeneity is crucial when our goal is not only characterising historical data, but also assessing the possible reductions in IFRs. For example, it can be argued that an HCT can use screening and provide medical care to achieve a rate of IFRs which is at least as low as the region or country with the lowest IFRs in our data^1^.

Although existing statistical packages for meta-analysis (both Bayesian and frequentist) could easily be used to model event rates such as IFR, (Więcek & Meager, 2020; Carpenter, 2016) these models may encounter problems or provide biased results when in a large proportion of studies no deaths are observed, as is sometimes the case for COVID-19 in younger populations. To address this, we use death data and estimates of prevalence as inputs instead of IFRs, so that the model is generative, following best practice for such models (Betancourt, 2016; Gelman et al., 2020). We then construct a new, reproducible model for IFRs. Methodological details of the model are described in the Appendix.^2^ Crucially, we assume that the fixed effect of age and random effects of location on IFR are on logistic scale, as is typical for meta-analysis models of binary data.

While our IFR estimates capture average risks within different age groups and even heterogeneity across regions or countries, they still refer to the general population (of a given age, in a given location). A prospective HCT participant would be screened for health issues and comorbidities, further reducing the risk in comparison to the members of general population. To account for this, we perform an additional analysis using OpenSAFELY, (Williamson et al., 2020) a large observational dataset on COVID-19 mortality factors that includes comorbidity and age information, as well as data on gender, comparing the risk in general population to lower-risk subpopulation^3^. Similarly to adjustment for heterogeneity across general populations, the adjustment for screening can be turned on or off in our tool.

Input data for the Bayesian IFR model is based on a non-systematic review of the literature and earlier meta-analyses, particularly by Levin *et al*, (Levin, Cochran, & Walsh, 2020) but we have also opportunistically included other studies, and data from other official sources as detailed in the appendix. However, the list of studies is not fixed, since new and better characterized datasets are becoming available over time. For this reason, we have made sure that incorporating additional prevalence and death data from newer studies and/or updated data sets is straightforward. We will continually update the model to assure that any estimates provided to participants or used for decisions include all relevant data, rather than only using data that was available when the analysis was first performed.

We use the age- and gender-specific data from Salje et al. (Salje et al., 2020) for the rate of death of hospitalized patients to impute the relative risk of hospitalization based on our meta-analysis for mortality risk.

Our model is available publicly, together with input data and source code for the tool and under an open license, at https://github.com/1DaySooner/RiskModel (Manheim, Więcek, Choi, & Wick, 2021).

### 2.2 Transforming individual risk into study risk

Once a suitable challenge virus is manufactured, itself a complex process (Catchpole et al., 2018), the risk of the individual from a challenge trial depends on the dose of virus given. The uncertainties about dose-response lead to a number of additional uncertainties about overall study risk. For other viruses, such as H1N1 and H3N2 influenza, a dose-response relationship has been found (Memoli et al., 2015; Han et al., 2019). The specific dose-response relationship, and its functional form must be determined experimentally, which is an outcome rather than an input of a dosing study like the one we are considering.^4^. This uncertainty is a key issue, so, as suggested by Morgan and Henrion, we advise that this structural uncertainty should not be treated as a probabilistic variable, and instead sensitivity analysis should be used to enable the consideration of a range of plausible outcomes (Morgan, Henrion, & Small, 1990).

Given the specifics of a study design, the relationship between individual risk and the risks in the overall study is straightforward, assuming independence of risk between individuals in the study.^5^ The risk to an individual of severe disease in the study given dose *d* is *S*_*d*_, and the risk of mortality is *M*_*d*_. The probability that someone in a group of size *N* experiences the corresponding outcome is 1 − (1 − *S*_*d*_) ^*N*^ and 1 − (1 − *M*_*d*_) ^*N*^. By simulating the probability of impact for each dosed group and, in the case of more complex studies, conditioning the trial of later groups on the results of earlier ones, we can find the overall risk in more complex studies.

In the current version we have restricted the tool to consider to a simple *N* -person study. This means the tool provides an upper bound for risk of mortality and hospitalisation, higher than in a more complex dose-escalation study^6^. However, the underlying model structure allows simulation of dose-dependent and/or conditional study designs, as necessary, to accommodate dose response information and allow for the simulation of more complex trials.

### 2.3 Interface

The web tool we built allows exploration of two related types of risk. The first allows individuals to explore their personal risk, for example, by gender and age, if they volunteer to participate in a study, while the second displays overall risk of a single round dosing study with a given number of participants. Assumptions on study design and the underlying risk can be adjusted to allow interactive exploration by policymakers and participants. In considering personal risk, the tool uses pre-calculated outputs from the risk model to calculate and display predictions for hospitalization rate and death rate for individuals as a function of age and sex.

Deaths are further translated into micromorts–the expected number of deaths per million events, a standard method of showing mortality risk often used for patient consent (Howard, 1980; Ahmad, Peterson, & Torella, 2015). Quantities which cannot be transformed into micromorts, are presented as probabilities.^7^

The tool allows both the public and policymakers to explore how overall risks change depending on differences in study design. This also helps maintain transparency into clinical trial design concerns, thereby better informing potential challenge trial volunteers. We also note that for trial designers and ethicists, the relationship between risk of different impacts and compensation is critical (Grimwade et al., 2020; Palmer & Schuck, 2020; Blumenthal-Barby & Ubel, 2020).

The importance of transparency and public engagement has been widely noted in the literature on challenge trials (Jamrozik & Selgelid, 2020a). For this reason, the tool is available for public use, and is already being used to inform people who have volunteered to be contacted to potentially participate in a challenge trial (1Day Sooner, 2020).

## 3 RESULTS

The analysis dataset for age-specific IFRs contains 167 data points from 34 studies, with each containing between 2 and 11 different age groups; all data are presented in Tables 3 and 4 of the Appendix and are included in the code repository accompanying this paper. A glance at available data also confirms the necessity of using a more complicated modelling approach: only 37 inputs contain individuals aged between 20 and 30; however, out of these 37, only three cover precisely the main age group of interest, that is, people between 20 and 30 years of age.

**TABLE 1.** Main parameters in the sensitivity analysis models using subsets of data.

| Included data | beta | sigma | tau | IFR, 20-29 year olds (per 1,000) |
| --- | --- | --- | --- | --- |
| All data (main model) | -8.80 (0.12) | 0.65 (0.09) | 0.15 (0.02) | 1.12 (0.01) |
| Median age between 10 and 75 | -8.72 (0.14) | 0.72 (0.11) | 0.17 (0.02) | 1.07 (0.01) |
| Median age over 10 | -8.79 (0.12) | 0.66 (0.09) | 0.15 (0.02) | 1.11 (0.01) |
| Median age under 75 | -8.75 (0.13) | 0.70 (0.11) | 0.16 (0.02) | 1.08 (0.01) |

**TABLE 2.**
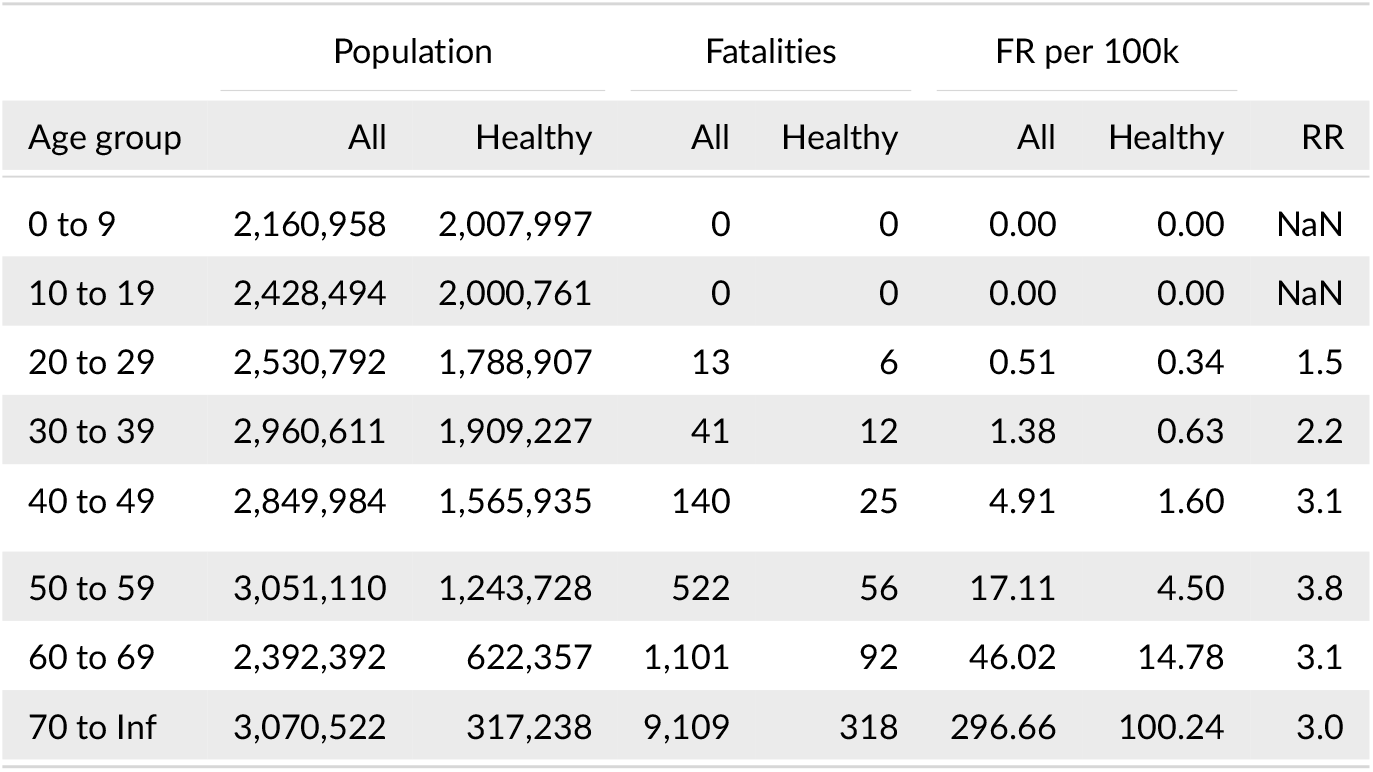
Data from the OpenSAFELY database grouped by age.

| Age group | Population |  | Fatalities |  | FR per 100k |  | RR |
| --- | --- | --- | --- | --- | --- | --- | --- |
|  | All | Healthy | All | Healthy | All | Healthy |  |
| 0 to 9 | 2,160,958 | 2,007,997 | 0 | 0 | 0.00 | 0.00 | NaN |
| 10 to 19 | 2,428,494 | 2,000,761 | 0 | 0 | 0.00 | 0.00 | NaN |
| 20 to 29 | 2,530,792 | 1,788,907 | 13 | 6 | 0.51 | 0.34 | 1.5 |
| 30 to 39 | 2,960,611 | 1,909,227 | 41 | 12 | 1.38 | 0.63 | 2.2 |
| 40 to 49 | 2,849,984 | 1,565,935 | 140 | 25 | 4.91 | 1.60 | 3.1 |
| 50 to 59 | 3,051,110 | 1,243,728 | 522 | 56 | 17.11 | 4.50 | 3.8 |
| 60 to 69 | 2,392,392 | 622,357 | 1,101 | 92 | 46.02 | 14.78 | 3.1 |
| 70 to Inf | 3,070,522 | 317,238 | 9,109 | 318 | 296.66 | 100.24 | 3.0 |

**TABLE 3.**
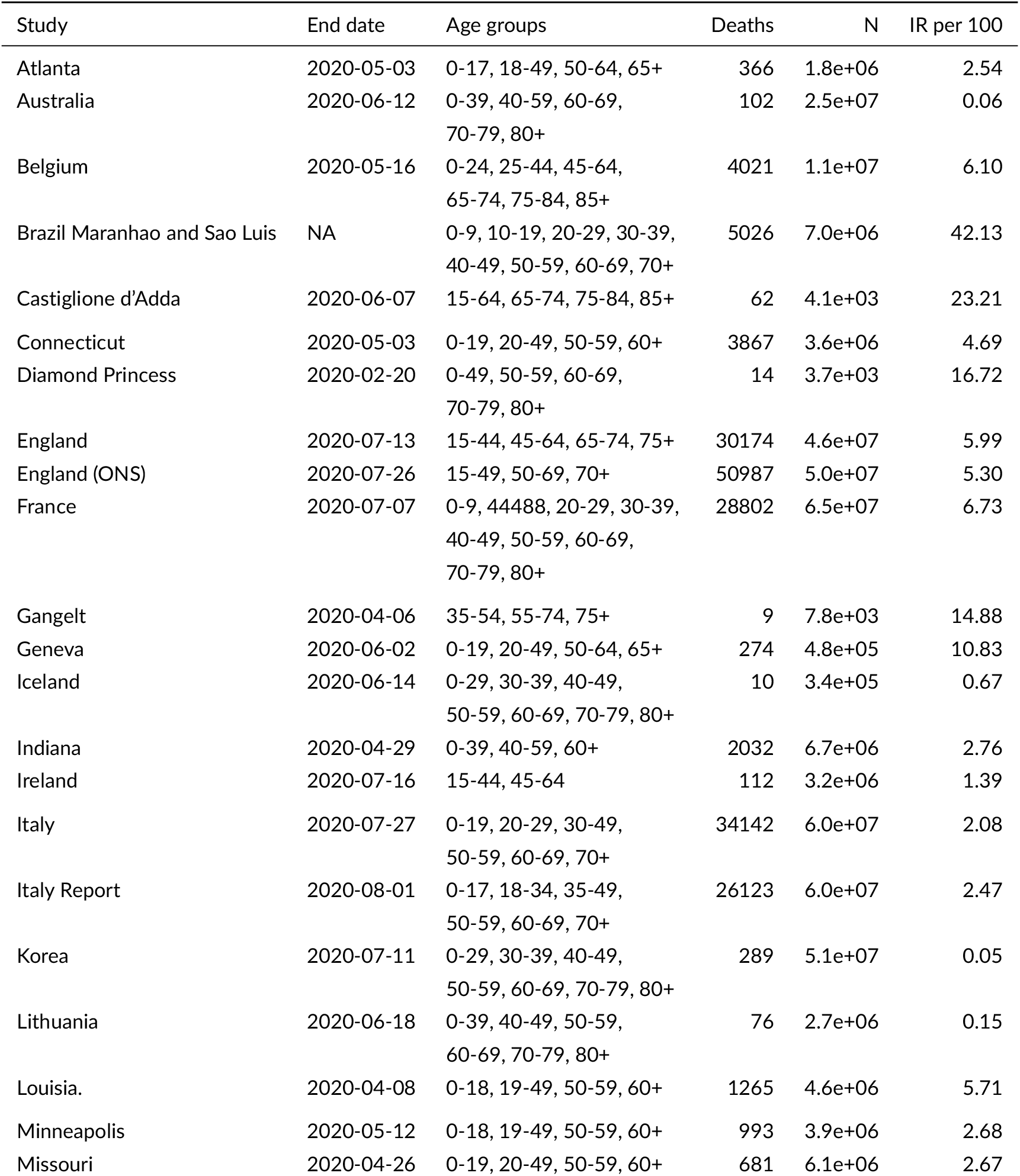
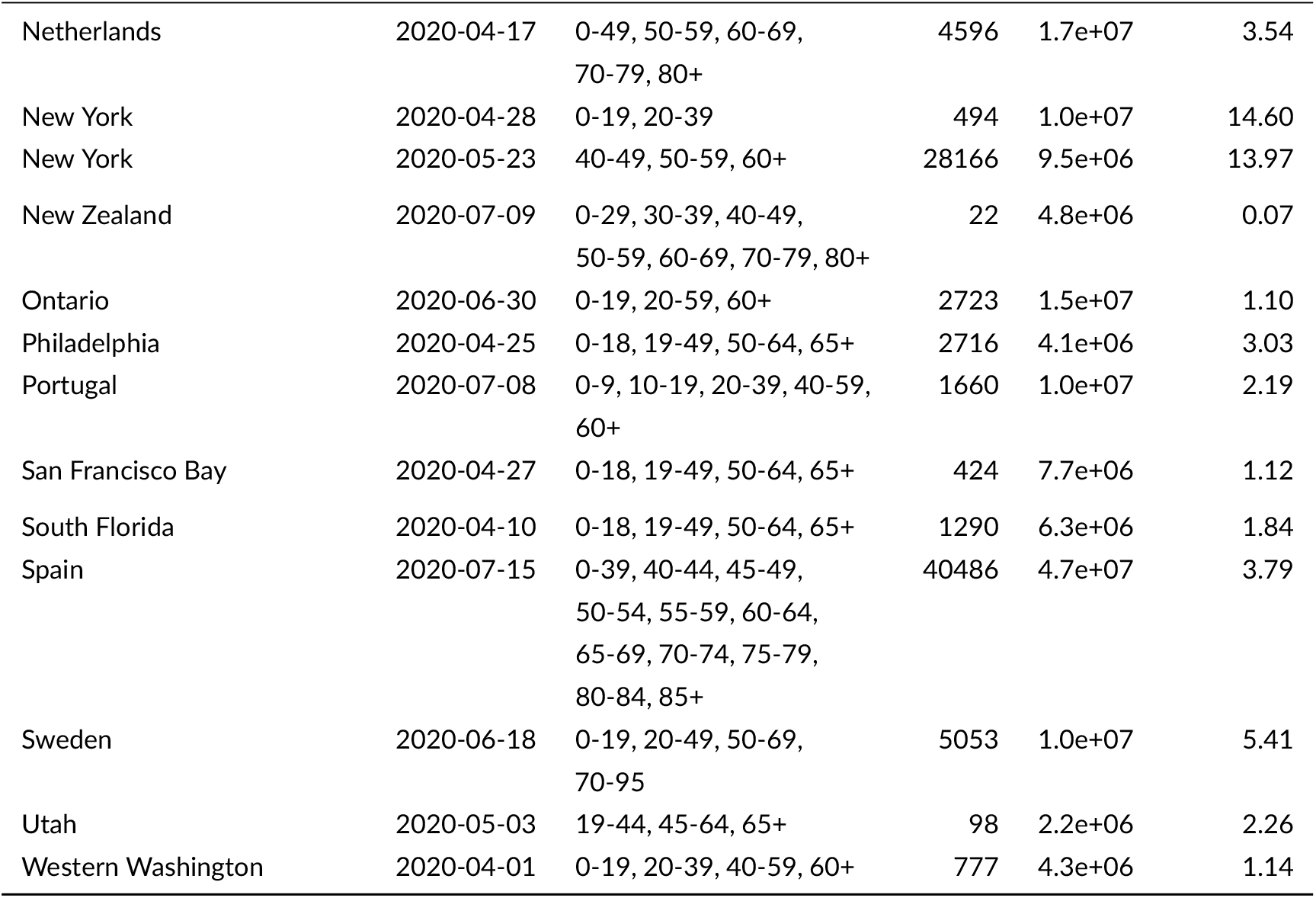
Complete table of studies used by the meta-analysis model.

| Study | End date | Age groups | Deaths | N | IR per 100 |
| --- | --- | --- | --- | --- | --- |
| Atlanta | 2020-05-03 | 0-17, 18-49, 50-64, 65+ | 366 | 1.8e+06 | 2.54 |
| Australia | 2020-06-12 | 0-39, 40-59, 60-69, 70-79, 80+ | 102 | 2.5e+07 | 0.06 |
| Belgium | 2020-05-16 | 0-24, 25-44, 45-64, 65-74, 75-84, 85+ | 4021 | 1.1e+07 | 6.10 |
| Brazil Maranhao and Sao Luis | NA | 0-9, 10-19, 20-29, 30-39, 40-49, 50-59, 60-69, 70+ | 5026 | 7.0e+06 | 42.13 |
| Castiglione d'Adda | 2020-06-07 | 15-64, 65-74, 75-84, 85+ | 62 | 4.1e+03 | 23.21 |
| Connecticut | 2020-05-03 | 0-19, 20-49, 50-59, 60+ | 3867 | 3.6e+06 | 4.69 |
| Diamond Princess | 2020-02-20 | 0-49, 50-59, 60-69, 70-79, 80+ | 14 | 3.7e+03 | 16.72 |
| England | 2020-07-13 | 15-44, 45-64, 65-74, 75+ | 30174 | 4.6e+07 | 5.99 |
| England (ONS) | 2020-07-26 | 15-49, 50-69, 70+ | 50987 | 5.0e+07 | 5.30 |
| France | 2020-07-07 | 0-9, 44488, 20-29, 30-39, 40-49, 50-59, 60-69, 70-79, 80+ | 28802 | 6.5e+07 | 6.73 |
| Gangelt | 2020-04-06 | 35-54, 55-74, 75+ | 9 | 7.8e+03 | 14.88 |
| Geneva | 2020-06-02 | 0-19, 20-49, 50-64, 65+ | 274 | 4.8e+05 | 10.83 |
| Iceland | 2020-06-14 | 0-29, 30-39, 40-49, 50-59, 60-69, 70-79, 80+ | 10 | 3.4e+05 | 0.67 |
| Indiana | 2020-04-29 | 0-39, 40-59, 60+ | 2032 | 6.7e+06 | 2.76 |
| Ireland | 2020-07-16 | 15-44, 45-64 | 112 | 3.2e+06 | 1.39 |
| Italy | 2020-07-27 | 0-19, 20-29, 30-49, 50-59, 60-69, 70+ | 34142 | 6.0e+07 | 2.08 |
| Italy Report | 2020-08-01 | 0-17, 18-34, 35-49, 50-59, 60-69, 70+ | 26123 | 6.0e+07 | 2.47 |
| Korea | 2020-07-11 | 0-29, 30-39, 40-49, 50-59, 60-69, 70-79, 80+ | 289 | 5.1e+07 | 0.05 |
| Lithuania | 2020-06-18 | 0-39, 40-49, 50-59, 60-69, 70-79, 80+ | 76 | 2.7e+06 | 0.15 |
| Louisia. | 2020-04-08 | 0-18, 19-49, 50-59, 60+ | 1265 | 4.6e+06 | 5.71 |
| Minneapolis | 2020-05-12 | 0-18, 19-49, 50-59, 60+ | 993 | 3.9e+06 | 2.68 |
| Missouri | 2020-04-26 | 0-19, 20-49, 50-59, 60+ | 681 | 6.1e+06 | 2.67 |

**TABLE 3** Complete table of studies used by the meta-analysis model. (*continued*)
| Study | End date | Age groups | Deaths | N | IR per 100 |
| --- | --- | --- | --- | --- | --- |
| Netherlands | 2020-04-17 | 0-49, 50-59, 60-69,<br>70-79, 80+ | 4596 | 1.7e+07 | 3.54 |
| New York | 2020-04-28 | 0-19, 20-39 | 494 | 1.0e+07 | 14.60 |
| New York | 2020-05-23 | 40-49, 50-59, 60+ | 28166 | 9.5e+06 | 13.97 |
| New Zealand | 2020-07-09 | 0-29, 30-39, 40-49,<br>50-59, 60-69, 70-79, 80+ | 22 | 4.8e+06 | 0.07 |
| Ontario | 2020-06-30 | 0-19, 20-59, 60+ | 2723 | 1.5e+07 | 1.10 |
| Philadelphia | 2020-04-25 | 0-18, 19-49, 50-64, 65+ | 2716 | 4.1e+06 | 3.03 |
| Portugal | 2020-07-08 | 0-9, 10-19, 20-39, 40-59,<br>60+ | 1660 | 1.0e+07 | 2.19 |
| San Francisco Bay | 2020-04-27 | 0-18, 19-49, 50-64, 65+ | 424 | 7.7e+06 | 1.12 |
| South Florida | 2020-04-10 | 0-18, 19-49, 50-64, 65+ | 1290 | 6.3e+06 | 1.84 |
| Spain | 2020-07-15 | 0-39, 40-44, 45-49,<br>50-54, 55-59, 60-64,<br>65-69, 70-74, 75-79,<br>80-84, 85+ | 40486 | 4.7e+07 | 3.79 |
| Sweden | 2020-06-18 | 0-19, 20-49, 50-69,<br>70-95 | 5053 | 1.0e+07 | 5.41 |
| Utah | 2020-05-03 | 19-44, 45-64, 65+ | 98 | 2.2e+06 | 2.26 |
| Western Washington | 2020-04-01 | 0-19, 20-39, 40-59, 60+ | 777 | 4.3e+06 | 1.14 |

**TABLE 4.** Complete table of inputs used by the meta-analysis model and crude IFR’s.

| Study location | Age | Deaths | N | Infection rate (%) |  |  | IFR/100k |
| --- | --- | --- | --- | --- | --- | --- | --- |
|  |  |  |  | Mean | 2.5% | 97.5% |  |
| Brazil Maranhao and Sao Luis | 0-9 | 29 | 1.2e+06 | 42.60 | 51.30 | 33.80 | 6.0e-02 |
| France | 0-9 | 3 | 7.5e+06 | 5.90 | 10.20 | 1.60 | 1.0e-02 |
| Portugal | 0-9 | 0 | 8.4e+05 | 2.20 | 6.00 | 0.90 | 0.0e+00 |
| Atlanta | 0-17 | 1 | 4.0e+05 | 0.00 | 1.00 | 0.00 | 1.0e+05 |
| Italy Report | 0-17 | 5 | 9.6e+06 | 2.20 | 2.80 | 1.70 | 2.0e-02 |
| Louisia. | 0-18 | 1 | 1.1e+06 | 2.80 | 11.50 | 0.00 | 3.0e-02 |
| Minneapolis | 0-18 | 0 | 9.7e+05 | 5.80 | 14.30 | 0.00 | 0.0e+00 |
| Philadelphia | 0-18 | 1 | 9.4e+05 | 2.20 | 6.90 | 0.00 | 5.0e-02 |
| San Francisco Bay | 0-18 | 0 | 1.6e+06 | 1.70 | 7.70 | 0.00 | 0.0e+00 |
| South Florida | 0-18 | 0 | 1.3e+06 | 2.40 | 7.80 | 0.00 | 0.0e+00 |
| Missouri | 0-19 | 0 | 1.5e+06 | 1.40 | 4.10 | 0.00 | 0.0e+00 |
| Italy | 0-19 | 4 | 1.1e+07 | 1.80 | 2.40 | 1.30 | 2.0e-02 |
| Connecticut | 0-19 | 2 | 8.3e+05 | 0.80 | 2.90 | 0.00 | 3.0e-01 |
| Geneva | 0-19 | 0 | 8.0e+04 | 9.17 | 14.06 | 5.40 | 0.0e+00 |
| New York | 0-19 | 12 | 4.9e+06 | 14.60 | 16.10 | 13.10 | 2.0e-02 |
| Ontario | 0-19 | 1 | 3.1e+06 | 0.80 | 1.40 | 0.30 | 4.0e-02 |
| Sweden | 0-19 | 1 | 2.3e+06 | 5.73 | 7.00 | 4.45 | 1.0e-02 |
| Western Washington | 0-19 | 0 | 1.0e+06 | 0.70 | 2.50 | 0.00 | 0.0e+00 |
| Belgium | 0-24 | 0 | 3.2e+06 | 6.00 | 8.60 | 4.20 | 0.0e+00 |
| Brazil Maranhao and Sao Luis | 10-19 | 21 | 1.3e+06 | 43.00 | 52.40 | 33.50 | 4.0e-02 |
| France | 44488 | 3 | 7.9e+06 | 3.50 | 6.40 | 0.70 | 1.0e-02 |
| Iceland | 0-29 | 0 | 1.4e+05 | 0.41 | 0.60 | 0.28 | 0.0e+00 |
| Korea | 0-29 | 0 | 1.6e+07 | 0.06 | 0.08 | 0.03 | 0.0e+00 |
| New Zealand | 0-29 | 0 | 1.9e+06 | 0.06 | 0.08 | 0.03 | 0.0e+00 |
| Portugal | 10-19 | 0 | 1.0e+06 | 2.40 | 6.10 | 0.90 | 0.0e+00 |
| Indiana | 0-39 | 20 | 3.5e+06 | 3.05 | 4.30 | 1.90 | 1.8e-01 |
| Spain | 0-39 | 225 | 1.9e+07 | 3.73 | 4.60 | 2.93 | 3.1e-01 |
| Australia | 0-39 | 0 | 1.4e+07 | 0.06 | 0.09 | 0.03 | 0.0e+00 |
| Lithuania | 0-39 | 0 | 1.2e+06 | 0.15 | 0.22 | 0.07 | 0.0e+00 |
| Italy | 20-29 | 16 | 6.2e+06 | 1.70 | 2.00 | 1.30 | 1.5e-01 |
| Brazil Maranhao and Sao Luis | 20-29 | 47 | 1.3e+06 | 49.20 | 57.30 | 41.10 | 8.0e-02 |
| France | 20-29 | 21 | 7.4e+06 | 7.00 | 10.20 | 3.80 | 4.0e-02 |
| Italy Report | 18-34 | 43 | 1.1e+07 | 2.10 | 2.40 | 1.70 | 1.9e-01 |
| Netherlands | 0-49 | 40 | 1.0e+07 | 3.50 | 5.20 | 2.50 | 1.1e-01 |
| England | 15-44 | 524 | 2.1e+07 | 7.20 | 7.66 | 6.73 | 3.4e-01 |
| Ireland | 15-44 | 18 | 2.0e+06 | 1.50 | 2.70 | 0.30 | 6.1e-01 |
| New York | 20-39 | 482 | 5.4e+06 | 14.60 | 16.10 | 13.10 | 6.1e-01 |

**TABLE 4** Complete table of inputs used by the meta-analysis model and crude IFR's. (continued)
| Study location | Age | Deaths | N | Mean | 2.5% | 97.5% | IFR/100k |
| --- | --- | --- | --- | --- | --- | --- | --- |
| Portugal | 20-39 | 4 | 2.3e+06 | 0.90 | 5.30 | 0.10 | 1.9e-01 |
| Western Washington | 20-39 | 8 | 1.3e+06 | 1.30 | 2.30 | 0.70 | 4.6e-01 |
| Philadelphia | 19-49 | 51 | 1.4e+06 | 5.90 | 9.80 | 2.40 | 6.0e-01 |
| Utah | 19-44 | 3 | 1.2e+06 | 1.80 | 3.50 | 0.60 | 1.4e-01 |
| England (ONS) | 15-49 | 1035 | 2.5e+07 | 6.10 | 7.50 | 4.90 | 6.8e-01 |
| Atlanta | 18-49 | 20 | 8.7e+05 | 3.30 | 6.40 | 1.60 | 7.0e-01 |
| Louisia. | 19-49 | 85 | 1.9e+06 | 7.40 | 10.00 | 4.70 | 6.1e-01 |
| Minneapolis | 19-49 | 18 | 1.6e+06 | 2.30 | 4.20 | 0.80 | 4.8e-01 |
| San Francisco Bay | 19-49 | 25 | 3.4e+06 | 1.10 | 2.60 | 0.00 | 6.7e-01 |
| South Florida | 19-49 | 61 | 2.5e+06 | 0.90 | 2.20 | 0.20 | 2.7e+00 |
| Missouri | 20-49 | 18 | 2.3e+06 | 3.40 | 5.50 | 1.40 | 2.3e-01 |
| Belgium | 25-44 | 18 | 3.0e+06 | 5.90 | 8.30 | 4.20 | 1.0e-01 |
| Brazil Maranhao and Sao Luis | 30-39 | 163 | 1.1e+06 | 44.40 | 51.40 | 37.40 | 3.4e-01 |
| Connecticut | 20-49 | 75 | 1.3e+06 | 6.10 | 9.30 | 3.10 | 9.2e-01 |
| Diamond Princess | 0-49 | 0 | 1.2e+03 | 8.26 | 8.28 | 8.24 | 0.0e+00 |
| France | 30-39 | 84 | 8.0e+06 | 3.40 | 5.80 | 1.00 | 3.1e-01 |
| Geneva | 20-49 | 1 | 2.2e+05 | 13.12 | 17.00 | 9.75 | 3.0e-02 |
| Iceland | 30-39 | 1 | 4.7e+04 | 1.00 | 1.50 | 0.70 | 2.1e+00 |
| Korea | 30-39 | 2 | 7.1e+06 | 0.04 | 0.07 | 0.02 | 6.4e-01 |
| New Zealand | 30-39 | 0 | 6.2e+05 | 0.08 | 0.12 | 0.04 | 0.0e+00 |
| Sweden | 20-49 | 63 | 3.9e+06 | 6.50 | 7.84 | 5.16 | 2.5e-01 |
| Ontario | 20-59 | 122 | 8.0e+06 | 1.00 | 1.30 | 0.70 | 1.5e+00 |
| Italy | 30-49 | 369 | 1.6e+07 | 2.00 | 2.40 | 1.70 | 1.1e+00 |
| Castiglione d'Adda | 15-64 | 4 | 3.1e+03 | 19.10 | 23.24 | 14.86 | 6.9e+00 |
| Italy Report | 35-49 | 334 | 1.3e+07 | 2.40 | 2.80 | 2.10 | 1.1e+00 |
| Spain | 40-44 | 78 | 4.0e+06 | 3.80 | 4.60 | 3.00 | 5.1e-01 |
| Brazil Maranhao and Sao Luis | 40-49 | 290 | 8.0e+05 | 32.20 | 41.00 | 23.40 | 1.1e+00 |
| France | 40-49 | 231 | 8.3e+06 | 7.70 | 10.90 | 4.60 | 3.6e-01 |
| Gangelt | 35-54 | 0 | 3.6e+03 | 14.00 | 18.00 | 12.00 | 0.0e+00 |
| Iceland | 40-49 | 0 | 4.3e+04 | 1.50 | 2.00 | 1.10 | 0.0e+00 |
| Korea | 40-49 | 3 | 8.2e+06 | 0.04 | 0.06 | 0.02 | 8.8e-01 |
| Lithuania | 40-49 | 1 | 3.6e+05 | 0.18 | 0.29 | 0.10 | 1.6e+00 |
| New York | 40-49 | 1026 | 2.4e+06 | 15.30 | 17.00 | 13.70 | 2.8e+00 |
| New Zealand | 40-49 | 0 | 5.9e+05 | 0.07 | 0.11 | 0.04 | 0.0e+00 |
| Spain | 45-49 | 196 | 3.9e+06 | 4.10 | 5.20 | 3.30 | 1.2e+00 |
| Indiana | 40-59 | 148 | 1.7e+06 | 3.14 | 5.00 | 1.90 | 2.8e+00 |
| Australia | 40-59 | 3 | 6.4e+06 | 0.06 | 0.10 | 0.04 | 7.8e-01 |
| Portugal | 40-59 | 75 | 3.1e+06 | 2.60 | 6.60 | 1.00 | 9.4e-01 |

**TABLE 4** Complete table of inputs used by the meta-analysis model and crude IFR's. (continued)
| Study location | Age | Deaths | N | Mean | 2.5% | 97.5% | IFR/100k |
| --- | --- | --- | --- | --- | --- | --- | --- |
| Western Washington | 40-59 | 69 | 1.1e+06 | 0.90 | 1.90 | 0.30 | 6.9e+00 |
| Spain | 50-54 | 230 | 3.6e+06 | 4.20 | 5.20 | 3.30 | 1.5e+00 |
| Italy | 50-59 | 1186 | 9.4e+06 | 2.70 | 3.10 | 2.30 | 4.7e+00 |
| England | 45-64 | 4657 | 1.4e+07 | 6.18 | 6.58 | 5.78 | 5.2e+00 |
| Louisia. | 50-59 | 126 | 5.9e+05 | 8.30 | 11.90 | 4.50 | 2.6e+00 |
| Minneapolis | 50-59 | 47 | 5.2e+05 | 0.70 | 2.80 | 0.00 | 1.3e+01 |
| Missouri | 50-59 | 43 | 8.0e+05 | 2.00 | 3.80 | 0.50 | 2.7e+00 |
| Philadelphia | 50-64 | 290 | 1.1e+06 | 0.80 | 2.80 | 0.00 | 3.4e+01 |
| Utah | 45-64 | 24 | 6.3e+05 | 2.90 | 5.20 | 0.90 | 1.3e+00 |
| Belgium | 45-64 | 280 | 3.1e+06 | 6.20 | 8.30 | 4.70 | 1.5e+00 |
| Brazil Maranhao and Sao Luis | 50-59 | 533 | 6.0e+05 | 39.10 | 46.10 | 32.10 | 2.3e+00 |
| Connecticut | 50-59 | 157 | 5.2e+05 | 8.10 | 11.60 | 4.80 | 3.7e+00 |
| Diamond Princess | 50-59 | 0 | 4.0e+02 | 14.82 | 14.90 | 14.70 | 0.0e+00 |
| France | 50-59 | 860 | 8.6e+06 | 9.70 | 13.10 | 6.40 | 1.0e+00 |
| Iceland | 50-59 | 0 | 4.2e+04 | 0.80 | 1.30 | 0.50 | 0.0e+00 |
| Ireland | 45-64 | 94 | 1.2e+06 | 1.20 | 2.10 | 0.30 | 6.4e+00 |
| Italy Report | 50-59 | 1196 | 9.6e+06 | 3.10 | 3.50 | 2.70 | 4.0e+00 |
| Korea | 50-59 | 15 | 8.5e+06 | 0.06 | 0.08 | 0.03 | 3.2e+00 |
| Lithuania | 50-59 | 3 | 4.2e+05 | 0.17 | 0.33 | 0.10 | 4.2e+00 |
| Netherlands | 50-59 | 137 | 2.5e+06 | 4.30 | 5.80 | 3.10 | 1.3e+00 |
| New York | 50-59 | 2764 | 2.6e+06 | 16.00 | 17.50 | 14.60 | 6.6e+00 |
| New Zealand | 50-59 | 0 | 6.3e+05 | 0.08 | 0.12 | 0.04 | 0.0e+00 |
| Atlanta | 50-64 | 51 | 3.3e+05 | 4.90 | 12.90 | 1.80 | 3.2e+00 |
| San Francisco Bay | 50-64 | 66 | 1.5e+06 | 0.70 | 2.40 | 0.00 | 6.4e+00 |
| South Florida | 50-64 | 169 | 1.3e+06 | 2.00 | 4.00 | 0.30 | 6.6e+00 |
| Geneva | 50-64 | 16 | 9.9e+04 | 10.45 | 14.11 | 7.31 | 1.6e+00 |
| Spain | 55-59 | 758 | 3.4e+06 | 3.90 | 4.90 | 3.10 | 5.7e+00 |
| England (ONS) | 50-69 | 7118 | 1.6e+07 | 4.80 | 5.80 | 4.00 | 9.4e+00 |
| Sweden | 50-69 | 504 | 2.4e+06 | 4.81 | 5.98 | 3.64 | 4.4e+00 |
| Spain | 60-64 | 1249 | 2.9e+06 | 3.50 | 4.60 | 2.70 | 1.2e+01 |
| Gangelt | 55-74 | 0 | 3.1e+03 | 17.00 | 23.00 | 12.00 | 0.0e+00 |
| Italy | 60-69 | 3433 | 7.3e+06 | 2.20 | 2.50 | 1.70 | 2.1e+01 |
| Australia | 60-69 | 13 | 2.7e+06 | 0.09 | 0.13 | 0.05 | 5.4e+00 |
| Brazil Maranhao and Sao Luis | 60-69 | 1155 | 3.9e+05 | 40.30 | 51.40 | 29.10 | 7.3e+00 |
| Diamond Princess | 60-69 | 1 | 9.2e+02 | 19.18 | 19.30 | 19.10 | 5.6e+00 |
| France | 60-69 | 2204 | 7.8e+06 | 10.00 | 13.50 | 6.50 | 2.8e+00 |
| Iceland | 60-69 | 2 | 3.8e+04 | 0.50 | 1.00 | 0.30 | 1.1e+01 |
| Italy Report | 60-69 | 3274 | 7.5e+06 | 2.60 | 2.90 | 2.10 | 1.7e+01 |

**TABLE 4** Complete table of inputs used by the meta-analysis model and crude IFR's. (continued)
| Study location | Age | Deaths | N | Mean | 2.5% | 97.5% | IFR/100k |
| --- | --- | --- | --- | --- | --- | --- | --- |
| Korea | 60-69 | 41 | 6.5e+06 | 0.05 | 0.08 | 0.03 | 1.2e+01 |
| Lithuania | 60-69 | 12 | 3.5e+05 | 0.13 | 0.20 | 0.08 | 2.6e+01 |
| Netherlands | 60-69 | 454 | 2.1e+06 | 3.50 | 5.00 | 2.50 | 6.1e+00 |
| New Zealand | 60-69 | 3 | 5.2e+05 | 0.07 | 0.10 | 0.04 | 8.3e+00 |
| Spain | 65-69 | 1905 | 2.4e+06 | 4.10 | 5.30 | 3.10 | 1.9e+01 |
| Castiglione d'Adda | 65-74 | 17 | 5.4e+02 | 31.30 | 37.30 | 25.40 | 1.0e+02 |
| England | 65-74 | 5663 | 5.6e+06 | 3.16 | 3.66 | 2.67 | 3.2e+01 |
| Belgium | 65-74 | 663 | 1.1e+06 | 4.10 | 7.20 | 2.30 | 1.4e+01 |
| Connecticut | 60+ | 3633 | 8.8e+05 | 4.20 | 6.00 | 2.30 | 9.9e+01 |
| Ontario | 60+ | 2600 | 3.4e+06 | 1.60 | 2.10 | 1.10 | 4.7e+01 |
| Indiana | 60+ | 1864 | 1.5e+06 | 1.65 | 2.40 | 1.00 | 7.5e+01 |
| Louisia. | 60+ | 1053 | 1.0e+06 | 4.40 | 8.00 | 1.50 | 2.3e+01 |
| Minneapolis | 60+ | 928 | 8.1e+05 | 1.00 | 3.20 | 0.00 | 1.1e+02 |
| Missouri | 60+ | 620 | 1.5e+06 | 3.20 | 4.60 | 1.90 | 1.3e+01 |
| Spain | 70-74 | 3230 | 2.2e+06 | 3.80 | 5.10 | 2.80 | 3.9e+01 |
| Western Washington | 60+ | 700 | 8.7e+05 | 1.70 | 2.70 | 0.90 | 4.7e+01 |
| New York | 60+ | 24376 | 4.5e+06 | 12.10 | 13.10 | 11.20 | 4.4e+01 |
| Portugal | 60+ | 1581 | 3.0e+06 | 2.70 | 5.40 | 1.20 | 2.0e+01 |
| Australia | 70-79 | 31 | 1.8e+06 | 0.08 | 0.12 | 0.04 | 2.1e+01 |
| France | 70-79 | 5650 | 5.7e+06 | 5.90 | 8.70 | 3.10 | 1.7e+01 |
| Geneva | 65+ | 257 | 8.4e+04 | 6.82 | 10.53 | 3.83 | 4.5e+01 |
| Netherlands | 70-79 | 1539 | 1.6e+06 | 3.00 | 5.30 | 1.70 | 3.2e+01 |
| New Zealand | 70-79 | 7 | 3.6e+05 | 0.04 | 0.07 | 0.02 | 4.4e+01 |
| Atlanta | 65+ | 294 | 2.3e+05 | 0.70 | 4.50 | 0.10 | 1.9e+02 |
| Diamond Princess | 70-79 | 8 | 1.0e+03 | 23.05 | 23.20 | 23.00 | 3.5e+01 |
| Iceland | 70-79 | 3 | 2.3e+04 | 0.30 | 1.30 | 0.27 | 4.3e+01 |
| Korea | 70-79 | 84 | 3.6e+06 | 0.05 | 0.07 | 0.03 | 4.8e+01 |
| Lithuania | 70-79 | 23 | 2.2e+05 | 0.09 | 0.14 | 0.05 | 1.1e+02 |
| Philadelphia | 65+ | 2374 | 6.8e+05 | 1.60 | 3.50 | 0.30 | 2.2e+02 |
| San Francisco Bay | 65+ | 333 | 1.2e+06 | 0.90 | 2.50 | 0.20 | 3.0e+01 |
| Utah | 65+ | 71 | 3.7e+05 | 2.70 | 5.00 | 0.90 | 7.2e+00 |
| South Florida | 65+ | 1060 | 1.2e+06 | 3.00 | 4.50 | 1.70 | 2.9e+01 |
| Spain | 75-79 | 6175 | 1.8e+06 | 3.40 | 5.00 | 2.40 | 1.0e+02 |
| England (ONS) | 70+ | 42834 | 8.8e+06 | 3.90 | 5.20 | 3.00 | 1.3e+02 |
| Sweden | 70-95 | 4485 | 1.5e+06 | 3.12 | 4.12 | 2.13 | 9.4e+01 |
| Italy | 70+ | 29134 | 1.0e+07 | 2.10 | 2.50 | 1.70 | 1.3e+02 |
| Castiglione d'Adda | 75-84 | 25 | 4.0e+02 | 36.60 | 44.90 | 28.30 | 1.7e+02 |
| Belgium | 75-84 | 1182 | 6.9e+05 | 7.00 | 11.70 | 4.20 | 2.4e+01 |

**TABLE 4** Complete table of inputs used by the meta-analysis model and crude IFR's. (continued)
| Study location | Age | Deaths | N | Mean | 2.5% | 97.5% | IFR/100k |
| --- | --- | --- | --- | --- | --- | --- | --- |
| Brazil Maranhao and Sao Luis | 70+ | 2788 | 3.4e+05 | 34.30 | 42.90 | 25.70 | 2.4e+01 |
| England | 75+ | 19330 | 4.8e+06 | 3.30 | 4.08 | 2.53 | 1.2e+02 |
| Gangelt | 75+ | 9 | 1.2e+03 | 12.00 | 27.00 | 6.00 | 6.4e+01 |
| Italy Report | 70+ | 21271 | 1.1e+07 | 2.50 | 2.90 | 2.10 | 8.1e+01 |
| Spain | 80-84 | 5192 | 1.3e+06 | 3.90 | 6.10 | 2.40 | 1.0e+02 |
| Diamond Princess | 80+ | 5 | 2.2e+02 | 25.00 | 25.10 | 24.90 | 8.6e+01 |
| Iceland | 80+ | 4 | 1.3e+04 | 0.20 | 2.50 | 0.10 | 1.6e+02 |
| Korea | 80+ | 144 | 1.9e+06 | 0.06 | 0.09 | 0.03 | 1.3e+02 |
| New Zealand | 80+ | 12 | 1.9e+05 | 0.04 | 0.06 | 0.02 | 1.5e+02 |
| Lithuania | 80+ | 37 | 1.7e+05 | 0.13 | 0.19 | 0.07 | 1.7e+02 |
| Netherlands | 80+ | 2426 | 8.4e+05 | 2.80 | 7.30 | 0.90 | 1.0e+02 |
| Australia | 80+ | 55 | 1.1e+06 | 0.05 | 0.07 | 0.03 | 1.0e+02 |
| France | 80+ | 19746 | 4.0e+06 | 7.30 | 10.30 | 4.20 | 6.7e+01 |
| Spain | 85+ | 21248 | 1.6e+06 | 2.85 | 5.60 | 1.56 | 4.6e+02 |
| Castiglione d'Adda | 85+ | 16 | 1.5e+02 | 42.10 | 53.10 | 31.10 | 2.6e+02 |
| Belgium | 85+ | 1878 | 3.3e+05 | 13.20 | 19.60 | 8.90 | 4.4e+01 |

The basic evidence synthesis model, which includes all data, but does not adjust for health status, finds that average IFR in 20-29 age group for the studies included in this analysis is 15.1 per 100,000 cases (95% uncertainty interval^8^ 11.8 to 19.2). Extending the HCT population to include 20-39 year olds gives mean IFR of 26.5 per 100,000 (95% interval 20.6 to 33.5).

The model assumes a log-linear relationship between age and IFR. As seen in Figure 1, the assumption is met across the entire age range we considered, from children to people older than 80. The model estimates that IFR increases on average 3.06-fold per each additional decade of age.

**FIGURE 1.**
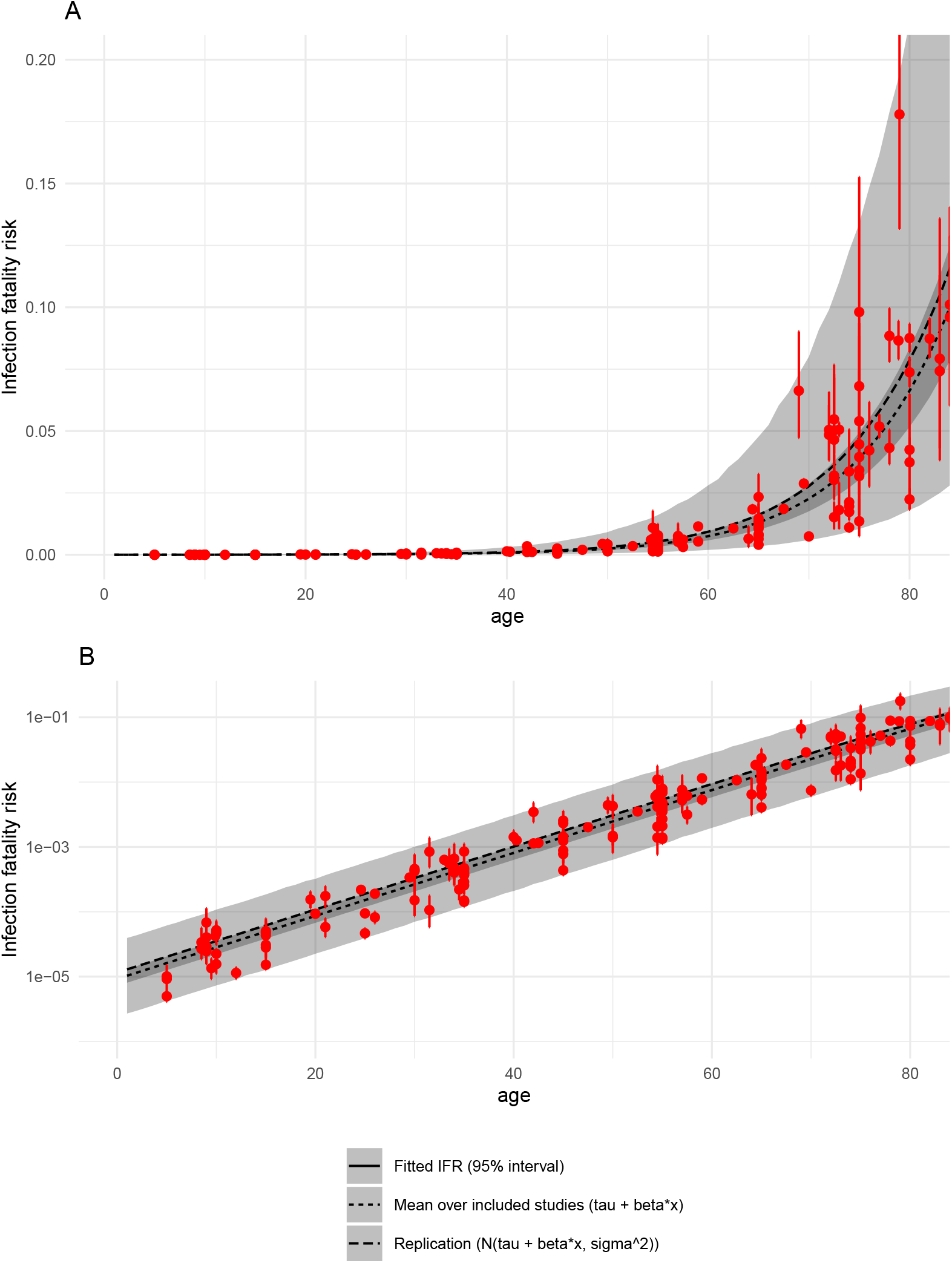
IFR as a function of age. Panel A is untransformed data. Panel B shows the same data on log 10 scale. Red points are data, *i.e*. model estimates of mean IFRs in particular studies, with bars representing 95% uncertainty intervals. Black and gray are modelled IFRs: lines are means, ribbons are 95% intervals: the narrower is average across all included studies, the wider takes into account heterogeneity between studies. Details and input values are given in Appendix.

These results align with two recent meta-analyses of IFRs (O’Driscoll et al., 2020; Brazeau et al., 2020). Both studies also show log-linear relationships between age and IFR, as well as considerable variation in IFR by context. O’Driscoll *et al* report median IFRs of 0.6 and 1.3 per 10,000 among those aged 20-24 and 25-29, respectively, while Brazeau *et al* report 3 and 4 per 10,000 respectively.

Based on OpenSAFELY data, we estimate that in healthy population (defined as lack of any co-morbidities listed above), the average mortality risk in 20-29 year olds is 1.9 times lower than in the general population, with 95% interval from 1.3 to 2.8. Expanding to 20-39 year olds only slightly increases the risk reduction factor, from 1.9 to 2. We also note that there is large heterogeneity across the studies, due to treatment availability and other factors^9^. The HCT volunteer population will receive the best available care, including the most up-to-date treatment options. They will also be screened based on socioeconomic and other population risk factors, trading off diversity for safety. For these reasons, we consider the population for an HCT to be akin to a best-case scenario. In the current analysis, this is France, where the estimated risk for 20-29 year olds is 4.67 × 10^−5^, approximately 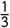 the mean risk across all populations.

It is unclear how much double-counting occurs when adjusting for both available medical care and population screening, and for screening out comorbidities. However, using this estimate, we see that a simple 50-person challenge trial has a 99.87% probability of having no induced fatalities, and a 98.9% probability of having no cases serious enough to require hospitalization. In other words, the probability of at least 1 death during a 50-person trial is 0.13% and the probability of at least 1 hospitalization is 1.1%. This represents an upper bound of risk for the notional 50-person trial, since the dosing study will escalate doses until a sufficient number of participants are infected, and therefore not everyone dosed is likely to develop an infection.

While the estimate is specific to a dosing study, it can also be useful for understanding the risk of later vaccine trials, though in that case other factors, including the possibility of vaccine-enhanced infection, would need to be assessed.

Due to lack of reliable data, the current model does not include estimates of longer term impacts, and instead includes a more qualitative discussion of these risks, (Smela, Kleinwaks, Sexton, & Schmit, 2020) but the model and interface will be updated with such estimates as they become available. The model interface can be used to explore how these uncertain factors can interact, as shown below in Figure 2.

**FIGURE 2.**
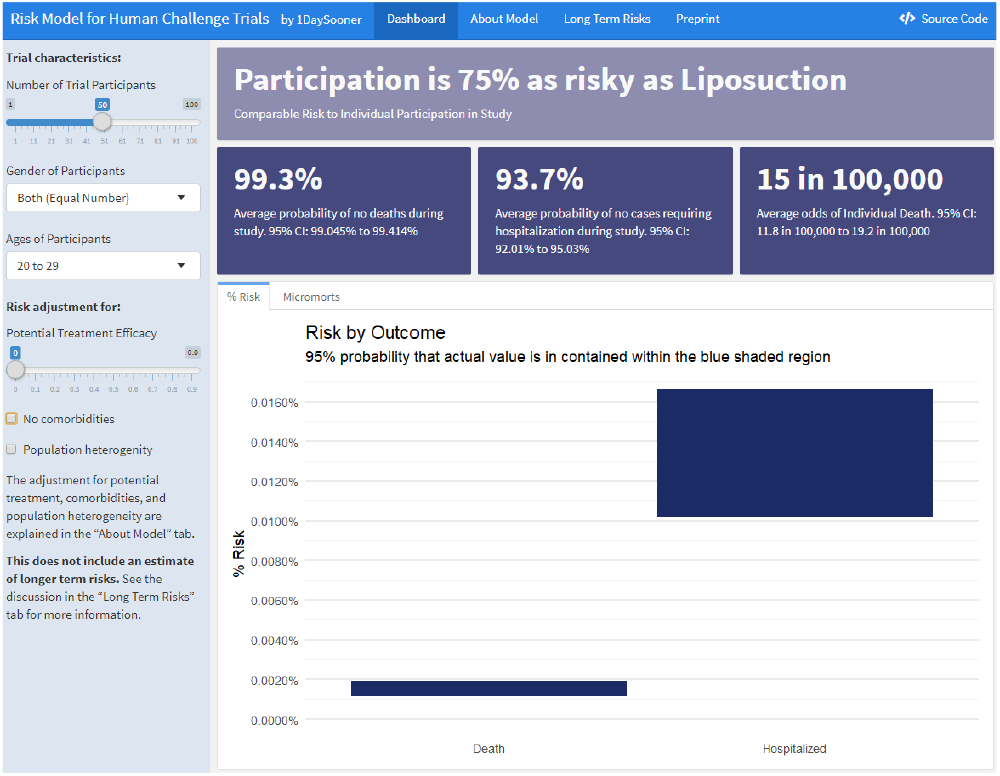
Interactive Model Interface

## 4 DISCUSSION

We have demonstrated that we can model certain risks associated with human challenge trials. However, our model is incomplete, and considerable concerns remain about the risks of human challenge trials. While a full accounting of challenge trial ethics is beyond the scope of this paper, we consider several factors below that inform our work on human challenge trial design, especially unmodeled risks. For a more complete perspective on the ethics of COVID-19 HCTs, we direct the reader to the World Health Organizations’ (WHO) key considerations (Jamrozik et al., 2020) and the recent discussion in Lancet Infectious Disease (Jamrozik & Selgelid, 2020a; Manheim, 2020) on how these issues are being addressed.

Our model suggests that the risk of a dosage-response study is far lower than other risks that are typically widely viewed as acceptable. The risk for participants selected to be low risk is clearly far lower than the risk in the general population from comparable clinical infections. While modelling the difference between the two is not possible, we are confident that the risk in HCT would indeed be lower than in observational, general population data that we used, due to more intense monitoring and availability of immediate high-priority medical care and pharmaceutical treatments, which we did not model here. We also note that HCTs have had historical precedent, showing promise for both less lethal human coronaviruses, and for yellow fever (Shah et al., 2017). They also provided early indications regarding the possible efficacy of a leading malaria vaccine candidate (Nielsen et al., 2018). The discussions about earlier trials shows that the ethical challenges of HCTs should not be seen as unique, but rather as laying along a natural continuum of clinical studies (Franklin & Grady, 2001). That said, the initial human challenge trials should be held to high ethical standards, both for individual risks and to preserve public trust in scientific and medical progress. That includes emphasis on fully informed consent of the participants, and, as noted by the WHO, higher than typical ethical standards (Jamrozik et al., 2020).

### 4.1 Limitations

Our model has a number of limitations. First, hospitalizations and deaths for 20-30 year olds are rare and may be subject to reporting bias. Second, we do not yet have enough information on long-term damage caused by COVID-19 and our model does not currently account for that risk, something we will discuss below. Third, although our model uses hospitalization as a proxy for the upper bound of serious non-fatal COVID-19 cases, more data are required to see if this is an accurate assumption.

Our model also may not capture changes in COVID-19 risks over time. It also does not estimate any indirect risks of the study. The model focuses on the risk due to the strain which will be used in an initial challenge trial. The model may not be appropriate for characteristing risk newer variants, including B.117. We stress that our model is not a comprehensive analysis of all available risks, but rather a tool quantifying certain known risks that can be used by trial participants and policymakers. We also limit our investigation to “risk” understood as probability of an event, as it is used in the medical literature(Kelly & Cowling, 2013), rather than a broader and more sociological understanding of risk.

Finally, this model captures only absolute risk. Relative risk assessment, comparing the risk of HCTs to other methods of finding the same information, is not intended. Any relative risk assessment would need to be combined with an assessment of relative benefits of each design, including speed and accuracy, as well as considering the ethical benefits of HCTs like the voluntary nature and better understanding and control of risks due to disease compared to other forms of clinical trials.

### 4.2 Opportunity Cost

When evaluating clinical trial designs, it is not sufficient to evaluate whether the proposed model is good, or even whether benefits outweigh costs, but also whether the alternatives are better. The key benefit of a dosing study is to allow further research with challenge trials, and alternative clinical trial models have major practical difficulties and far higher costs in a variety of ways. While this is not relevant for the initial set of vaccines that have now been approved, the challenge of conducting large scale trials is magnified for later vaccines.

For example, typical clinical phase 3 efficacy trials for an ongoing novel pandemic would be field trials, which rely on high numbers of trial participants, and require that a large number of people are treated with a new vaccine or drug. Such trials are relatively expensive and expose more trial participants to negative side effects of a given treatment, so that in many scenarios HCTs have been shown to be superior (Berry et al., 2020). These trials are also difficult to pursue now that an initial vaccine is available, due to ethical and logistical constraints. At this point, it may not be possible to find enough willing participants for field trials to study additional, potentially more effective or safer, vaccines.

### 4.3 Non-modeled Risks

We also note that there are several impacts we do not model in the study, most notably the concern about so-called “long COVID”, which is a catch-all term referring to a combination of persistent symptoms, slow recovery, and new post-recovery symptoms (Carfì, Bernabei, Landi, & Against COVID-19 Post-Acute Care Study Group, 2020). It is understood that for some cases, especially severe ones, recovery from COVID-19 can take months. In other cases there are longer-term symptoms differing from those experienced during the infection, perhaps similar to Post-SARS syndrome (Perrin et al., 2020; Moldofsky & Patcai, 2011). At the same time, COVID-19 recovery has been found to be faster in younger, healthier patients (Tenforde et al., 2020), which may mean the risk is lower in this group. It seems clear that the risk is a subject of continuing scientific investigation, and as it becomes better understood and quantified, it will be incorporated into both the risk model, and the web tool. Until then, the model contains an embedded overview of what is currently understood and known or unknown about longer term risks (Smela et al., 2020).

We also note that vaccine-induced disease enhancement is a critical concern for vaccine challenge trials, but is not relevant to dosing studies. Still, this risk must be considered in analyses of risks for later trials, and the current model would need to be adapted or supplemented to consider this if it were used to inform decisions regarding those later trials.

## 5 CONCLUSION

Human challenge trials are not risk free, but the balance of risk and benefits seems to clearly favor allowing them, as a large group of experts has argued (Sooner et al., 2020). This conclusion is disputed by some, but all decisions are made despite uncertainties and debate – whether empirical or moral (Lockhart, 2000; MacAskill, Bykvist, & Ord, 2020). The question is whether the both empirical and moral balance of factors lead to the indicated conclusion. The alternative is a failure to act due to misguided risk-aversion, or worse, using uncertainty and disputed moral claims as a positive stance to shut down further work, as has occurred in the debate about HCTs (Martinez et al., 2020).

It seems likely that HCTs are a viable way to rapidly test vaccine efficacy, which is particularly critical now for testing second-generation vaccines, which are important in quickly expanding vaccine availability (Castillo et al., 2021) and may prove superior to first generation vaccines. A dosing study is an urgent first step, and the risk estimates and tools developed for this paper can assist in planning such studies and informing volunteers.

Our model provides insight into the overall risk of a trial of a given size, and can better inform HCT participants about the dangers they face. Given that an HCT may help select the multiple vaccines necessary for global immunization while also assisting with therapeutic testing, the risk of an initial study into SARS-CoV-2 pathogenesis seems justified. If the dosing study is successful, future HCTs of COVID-19 may provide a rapid and systematic way of screening vaccine candidates for efficacy and safety, which is a significant benefit.

The results presented here are a useful static estimate of risk. The model is already being used to inform potential volunteers (1Day Sooner, 2020) and can be adapted and expanded in the future. Given the evolving understanding of the disease, the model should be continually updated with additional data on mortality and hospitalisations risks or other long-term risks. This will contribute to the discussion of whether or not to pursue challenge trials, which can help with response to COVID-19 in a variety of ways (Nguyen et al., 2020). Challenge trials may be an important tool for fighting COVID-19 and our model is a step towards that goal.

## Data Availability

All data and code is available on github, as linked in the paper.

https://github.com/1DaySooner/RiskModel

## Abbreviations

HCT: Human Challenge Trial
IFR: infection fatality risk
SARS-CoV-2: severe acute respiratory syndrome coronavirus-2

## Acknowledgements and thanks

Thank you to Rouslan Karimov and Jupiter Adams-Phipps for validating, updating, and collecting references for the dataset used in the analysis. Thanks to Chris Choe, Sophie Rose, Kate Sheahan, Troy Yamaguchi and Linchuan Zhang, who helped prepare and edit an early version of the manuscript. We would like to thank Ed Choi and William Kahn, as well as two anonymous referees, for providing helpful comments, and Ben Goldacre and the OpenSAFELY team (https://opensafely.org/team/) for sharing data with us.

## Appendix: Bayesian model of COVID-19 mortality risk in HCT volunteers

### A-1 INTRODUCTION

This short document is a technical appendix to the paper discussing COVID-19 risks in human challenge trial. Here, we show how a Bayesian model can synthesise information on many infection fatality risks (IFRs) into a single estimate. This estimate is specific to certain age groups and can be further adjusted by e.g. co-morbidity status. The analysis presented here is a form of Bayesian meta-analysis, in that our primary objective is to weigh sources of evidence in a way that captures both variability (here, heterogeneity in real IFRs across different settings) and uncertainty (here, the fact that we do not know the IFRs in each setting precisely).

The ultimate objective of this model is to characterise risk in a way that is useful for design of HCTs. Therefore, as a minimum, we want to incorporate variability across different populations into our prediction. Even better would be to understand how different factors can drive heterogeneity: *a priori* we hypothesise that the three main drivers of differences in IFRs are time-specific, population-specific and otherwise country-specific.^A1^

To characterise differences in observed IFRs we first develop a Bayesian model and apply it to publicly available summary data on IFRs from multiple countries and contexts, with particular focus on the impact of age. This is covered by Section 2. We then use a simple model to hypothesise reduction in risk that may be achieved by screening individuals for comorbidities; this is Section 3. We summarise all results in Section 4.

### A-2 AGE-SPECIFIC RISK OF COVID-19 MORTALITY

#### A-2.1 Bayesian evidence synthesis model

What follows is an adaptation of typical methods of Bayesian evidence synthesis to analysis of IFRs. IFR is the ratio of deaths to infections in a given population. Early estimates of COVID-19 mortality risk, e.g. by Verity, Okell, et al. (2020a), placed it at over 0.6%; however, it was also evident from data that IFR could be orders of magnitude higher in particular high risk groups, especially in the elderly, than in the general population.^A2^

By definition, our data on IFRs is a combination of data on deaths with data on infections. Typically, these are disjoint samples, in that numbers of infections are estimated (typically very imprecisely) on select subpopulation, while deaths are recorded in the general population (at a level of country, administrative region etc.). There are clear reasons to believe that IFRs will differ across studies (e.g. due to age, comorbidity status, time, genetic factors, quality of healthcare etc.). To address this, we will use a Bayesian hierarchical modelling framework to assume that the setting-specific estimates of *IF R*_*k*_ can differ from each other but are linked through some common parameters. (By *k* ‘s we denote different populations; note that sometimes we may have multiple *IF R* ‘s from different age groups in the same location.)

The most straight-forward and “canonical” way to implement such a Bayesian model is by modelling log odds of the event.^A3^ Deeks (2002) present a general treatment of such approach in medical statistics. Note, that for very rare events the odds of mortality are very similar to probability of mortality, but we model events on odds scale as a good “generic” approach to modelling binary data (in this case death following infections).^A4^

Basic models for this type of analysis of binary data can be implemented using existing statistical analysis packages; see, for example, *metafor* package in R or *baggr* by Więcek and Meager (2020). Such analysis would treat IFR as a logit-normal parameter to meta-analyse. However, note that when no deaths are observed, analysis of IFR (equal to observed deaths divided by modelled infections) is problematic. Therefore we propose a “custom” model that built in Stan which treats deaths and *prevalences* (rather than the IFRs) as data.

Let *d*_*k*_ denote observed deaths for data point *k* and assume that logit of corresponding prevalence estimate is *p*_*k*_ is a parameter (typically obtained from a statistical modelling papers, government reports etc.). Total population in *k* -th setting is *n*_*k*_. Total number of estimates is *K*. Then the model likelihood is as follows:

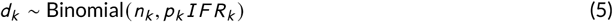

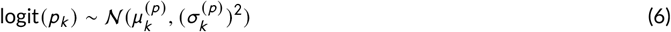

where 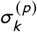 and 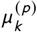 are parameters obtained from the literature (or converted from these parameters – see next section). The *k* data points collected can span many locations (studies); we denote them by loc_*k*_ and the total number of locations by *K*_*l oc*_ (with *K*_*l oc*_ < *K*).

In this model we can also account for various covariates impacting the IFRs (let’s denote their total number by *N*_*p*_), such as age groups (which we identify with median age of the population being studied, MedianAge_*k*_). We code them in a design matrix *X*. To center our *X* at the value of interest in our model (risk in 20-30 year olds), we use a transformation `MedianAge/10 - 2.5` to construct our matrix *X*. We denote all of the covariates using a design matrix *X* and denote by *N*_*p*_ the number of columns in *X*. We assume the impact on IFR is on logit scale, same as in the “canonical” logistic models of binary data that we mentioned above:

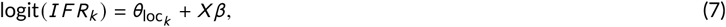

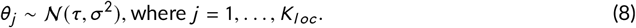

This means *θ* spans location-specific (random) effects on IFR while *β* is *N*_*p*_ dimensional vector of (fixed) covariate effects.

We implement our model in Stan and assume very weakly informative priors on all parameters, with prior for *τ* centered at 1 death per 10,000 cases.

~~~
 model {
 //Uncertain prevalence estimates (mu^p and sigma^p above):
 logit_prevalence ∼ normal(mean_prevalence, sd_prevalence);
 //Likelihood of mortality (d_k = obs_deaths, p_k = prevalence):
 obs_deaths ∼ binomial(population, prevalence .* ifr);
 //Hierarchical component of the model (location-specific theta):
 //logit_ifr = theta_k[loc] + to_vector(X*beta);
 theta_k ∼ normal(tau, sigma);
 //Priors:
 tau ∼ normal(logit(.0001), 5);
 sigma ∼ normal(0, 10);
 beta ∼ normal(0, 10);
 }
~~~

#### A-2.2 Data

We used estimates originally collected by Levin, Cochran, and Walsh (2020) to construct the first version of analysis dataset, which we then supplemented with more values extracted from other studies.

We included all relevant data points from the Levin et al. study This meant sometimes including values that the original study omitted. For example, Levin et al. exclude data points with seroprevalence indistinguishable from zero; our model retains them. We systematically went through the included studies listed in Appendix I and Appendices H.4 and H.5 of Levin et al. to ensure complete inclusion of all their data. In addition, in the course of this process we because aware of either updates to the studies Levin et al. used or entirely new relevant studies. We have reviewed and included them in our model as well.

The input data into our model consists of deaths (treated as known) and prevalences (treated as logit-distributed parameter with known mean and SD) in all reported age groups in all studies^A5^.

All of input data are given in Table 4. The analysis dataset contains 167 data points from 34 studies, each containing between 2 and 11 different age groups. We made only minimal modifications to source data, by 1) imputing the values from the Italian fatality data based on a seroprevalence survey, 2) imputing population size in Maranhao (as ratio of the reported number of infections and the mean infection risk) which were not reported and 3) assuming that uncertainty in prevalence 0-29 age group in Iceland is same as in the 30-39 age group since data were missing.

As mentioned, our model treats number of COVID-attributable deaths as measured without error (due to lack of data) but accounts for uncertainty in infection risks, which are always model-based estimates extracted from various available data sources. In preparing data, we assumed that logits of prevalence estimates from available studies are normally distributed, which seems to reproduce majority of data very well, see Figure 7 in the Supplement. There are some discrepancies with studies that allowed for prevalence estimates to be 0, something that our logit model does not allow.

For each study we construct a median age scalar defined by the average of the endpoints of each age range, rather than attempting to calculate a population-weighted mean.

Our approach of regressing on the median age and use of all available data (rather than the subset of data available in younger adults only) is necessitated by data limitations: out of 167 data points comprising age-specific estimates of prevalence (or IFR) and counts of deaths, 37 contain individuals aged 20-30 who are of primary interest to us. However, the populations are mixed with regards to age, with typical age groupings such as 19-49, 20-49, 20-39, 0-49 used instead. In fact, we find only one estimate out of 37 that is entirely specific to the 20-29 age group (Brazilian state of Maranhao), while one more has median age falling between 20 and 30 but is not specific to that age group.

#### A-2.3 Results

There were no issues with convergence of the Bayesian model. We set number of iterations to 5,000 and used 4 chains, with `max_treedepth` option set to 15. There were no divergent transitions and effective sample size was greater than 2310 for all of 538 modeled parameters (this number includes fitted prevalences, IFRs, *θ*’s and their transformations into/from logit scales). For the three main parameters in the model we obtained the following:

~~~
 ## Inference for Stan model: ifr_with0.
 ## 4 chains, each with iter=5000; warmup=2500; thin=1;
 ## post-warmup draws per chain=2500, total post-warmup draws=10000.
 ##
 ##          mean se_mean sd 2.5% 25% 50% 75% 98% n_eff Rhat
 ## tau      −8.80    0 0.12 −9.05    −8.88    −8.80    −8.72    −8.56    10954     1
 ## sigma     0.66    0 0.09 0.51     0.59     0.65     0.72     0.87     11075     1
 ## beta[1]   1.12    0 0.01 1.10     1.11     1.12     1.12     1.13     2556      1
 ##
 ## Samples were drawn using NUTS(diag_e) at Wed Feb 17 17:43:34 2021.
 ## For each parameter, n_eff is a crude measure of effective sample size,
 ## and Rhat is the potential scale reduction factor on split chains (at
 ## convergence, Rhat=1).
~~~

The mean coefficient of beta 1.12 corresponds to 3.06-fold increase in mortality risk following an infection per each extra decade of age (95% uncertainty interval is 3.01-3.11).

From these parameters we can predict average risks for subjects of any given age *x*, by using the posterior distribution of *τ* + (10*x* + 2.5)*β* (where 2.5 and 10 refer to the transformation that we applied to `MedianAge` inputs).

#### A-2.4 Average infection fatality risk in young subjects

Since we centered our `MedianAge` at 25 years in constructing our matrix *X*, we can now obtain model-estimated risk for a typical HCT population (aged 20 to 30, with median 25) by ignoring the *β* coefficient and examining *τ* and *σ* only. We find that the average IFR for this group (equal to 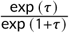) is 1.51 × 10^−4^ (with 95% interval from 1.18 × 10^−4^ to 1.92 × 10^−4^).

##### A-2.4.1 Heterogeneity in IFRs

However, there is a considerable variability in IFRs across different locations/dataset that we should consider. To take into account parameter *σ*, we can generate draws from the N(*τ, σ*^2^) distribution, corresponding to a hypothetical IFR in a new source of data. 95% interval for such model runs from 3.94 × 10^−5^ to 5.79 × 10^−4^. Since the model works a logistic scale, another way of interpreting the across-dataset variability is reporting the fold-impact of *σ* on the mean IFR; here, we obtain on average a 3.82-fold increase (decrease) in IFR per 2*σ* increase (decrease).

The lower end of the 95% interval, 3.94 × 10^−5^, is not extreme given input data, where the “crude” mean IFR (based on mean prevalence only) is below 7 per 10,000 for all data except for South Florida, and as low as 0 for some countries that did not record deaths (Belgium, New Zealand, Korea, Iceland) in various age groups including 20-29 year olds or 1.4 per 10,000 in Utah, in the population aged 19-44. (Please refer to Table 4 for complete list of inputs.)

We can assess this heterogeneity by inspecting the distribution of random effects in the model transformed into IFRs, i.e. the inverse logit transformation *θ* parameters. The largest (posterior mean) IFR value of *θ* is 5.09 × 10^−4^ in Castiglione d’Adda. The smallest posterior mean for 20-29 year olds is 4.67 × 10^−5^ in France.

##### A-2.4.2 Predictive checks for the model

We constructed posterior predictive distributions for number of deaths in each of the inputs by using the `generated quantities` functionality of Stan. Figure 3 compares the posterior means and 95% intervals with observed deaths. Out of 167 observations that were used to fit the model, 157 were within 95% intervals of the posterior predictive distributions. We observed the largest discrepancies occurred in Spanish data. Overall, we conclude that the simple binomial model we used here is flexible enough to capture both age-specific risk increases and heterogeneity in IFRs across settings/countries.

**FIGURE 3.**
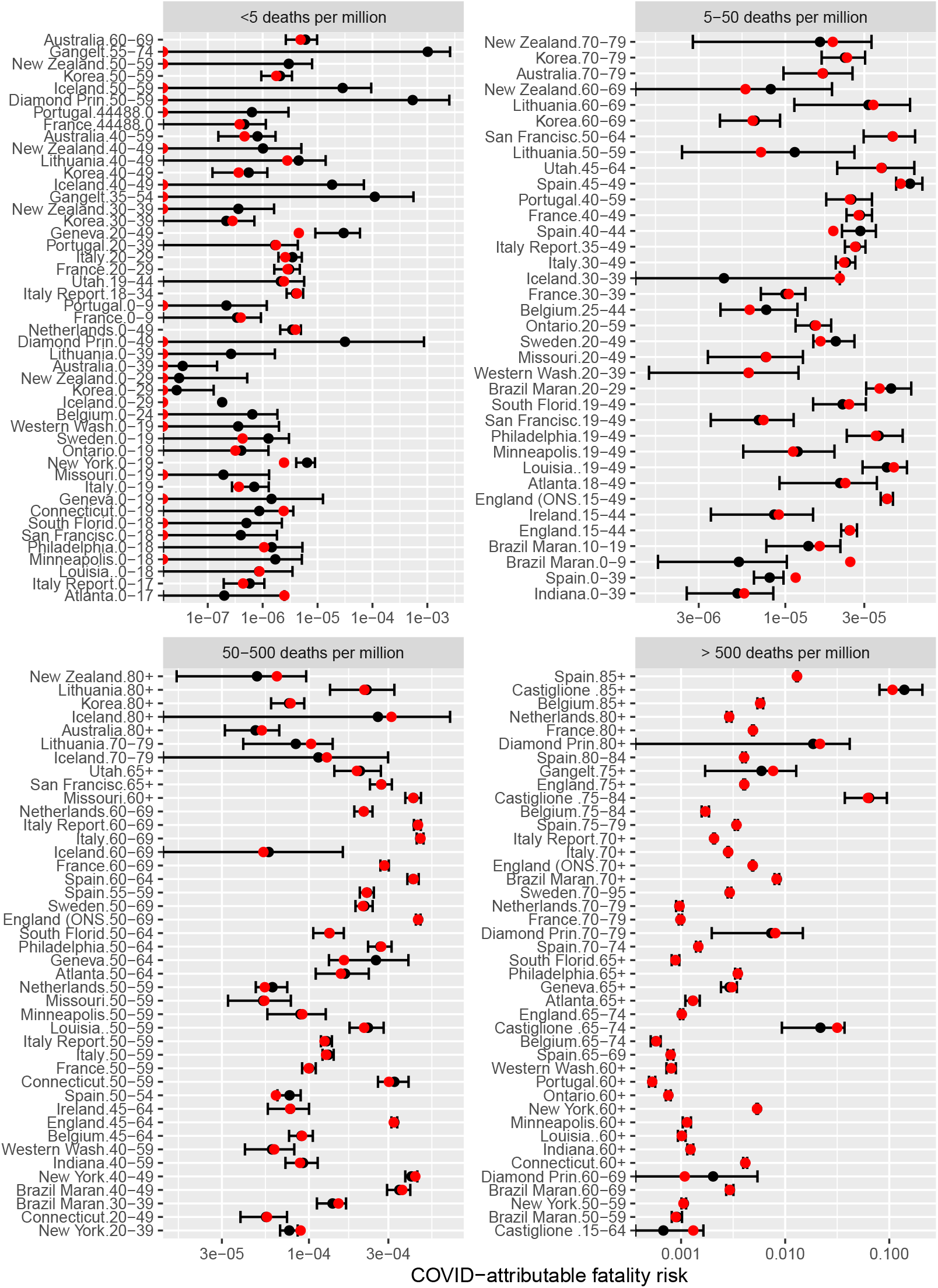
Comparison of model estimates (black) with data on observed fatality risk (FR, red), compared on logarithmic scale. FR is number of deaths divided by overall population size. Bars are 95% posterior interval; point is the mean. For better clarity, we grouped the plot into four panels according to observed FR X axes on each panel differ. For many low-risk populations (upper-left quadrant) no deaths were reported: we indicate this by plotting a red point on the left-hand side of the panel plot.

**FIGURE 4.**
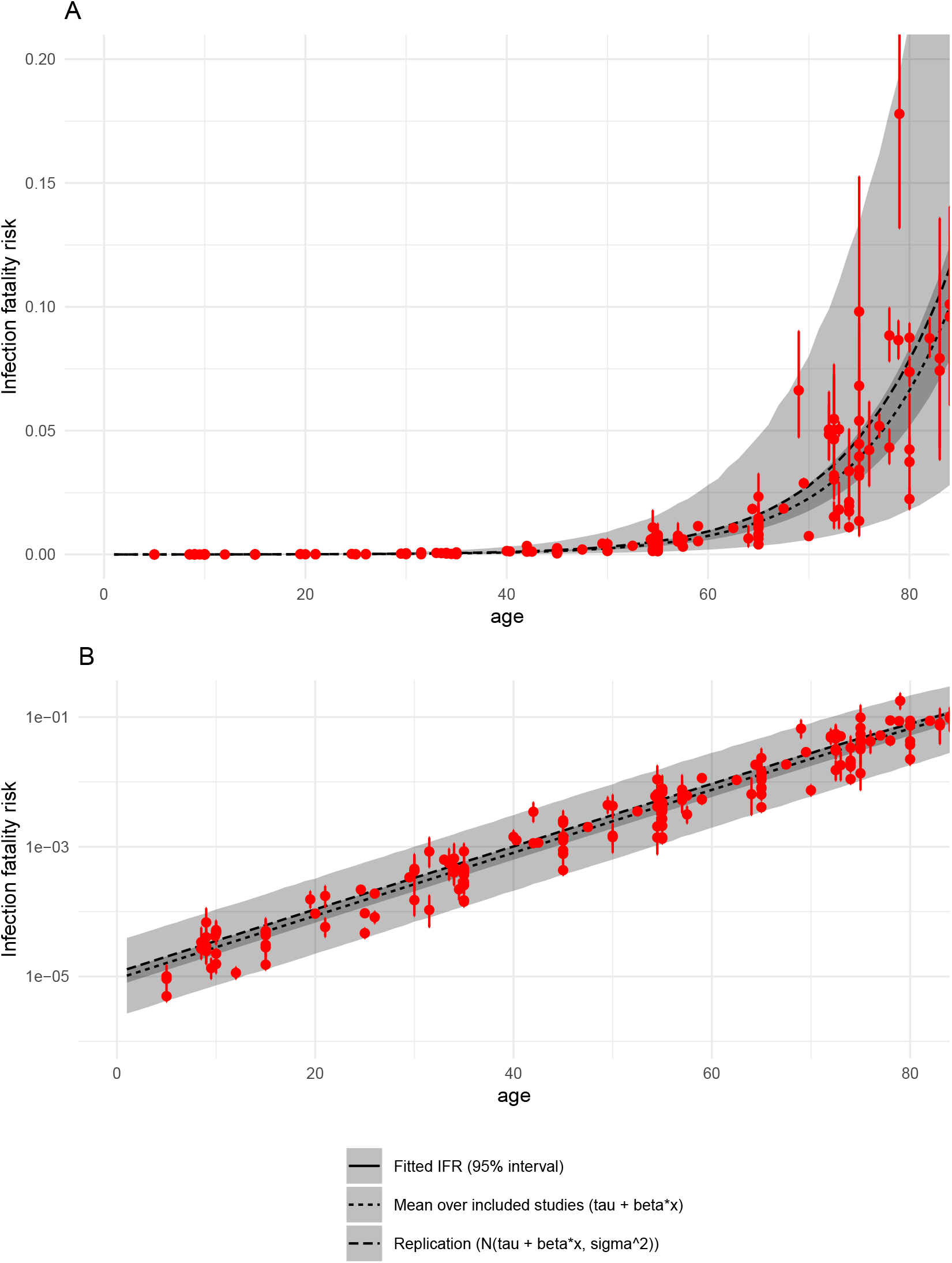
IFR as a function of age. Narrower ribbon corresponds to the 95% posterior interval of average across all included studies (tau parameter in the meta-analysis model), while the wider band takes into account heterogeneity (tau and sigma). Lines are means. Red points are model estimates of mean IFRs in partciular studies, with bars representing 95% posterior intervals. Panel A is untransformed data. Panel B shows the same data on log 10 scale.

#### A-2.5 Sensitivity analyses

As a sensitivity analysis, we also considered the impact on the main model parameters of dropping some data from our analysis. The result is summarised in a table containing parameters *σ, β, τ* and the mean IFR for the 20-29 year olds age group.

We considered excluding data for the youngest and the oldest individuals, as well as excluding both at once. Our hypothesis was that at extreme ends of age the assumption of log-linearity of IFRs may not hold and potentially lead to a biased estimate of the IFR in the 20-29 and 20-39 age groups. However, as shown in the accompanying table, we find no substantial effect of excluding data on the main model paramters.

### A-3 RISK REDUCTION IN HEALTHY INDIVIDUALS

We now turn our attention to the question of how much a human challenge trial designer could reduce the mortality risk by using simple screening methods, as discussed in the main paper.

Data for this section has been provided by OpenSAFELY (https://opensafely.org/) and was used by Williamson et al. (2020) to characterise COVID-19 mortality risk factors for 10,926 COVID-19 deaths in England. We group the total of 21,444,863 individuals into a total population and a lower-risk sub-population, defined as non-smoker, non-obese and without the comoribidities reported in the OpenSAFELY study^A6^, most notably respiratory and cardiovascular diseases and type I diabetes. For brevity we refer to the population without one of the pre-defined comorbidities as “healthy”. In contrast to the cited publication, we include records of individuals under 18 in our assessment. Counts grouped by age are presented in Table 2. Complete data (broken down by gender) are in Table 2 at the end of the document.

As shown in Table 2, for the age group of 20-29 the crude risk ratio (of general population vs the healthy subset only) is 1.53, but, due to low number of events in both healthy and general population, with a very wide 95% interval from 0.6 to 5.65.^A7^ As data on relative risks in other age groups is clearly related to the relative risk in 20-29 age group, we use another meta-analysis model to improve our estimate. Additionally, relative risks are higher in women than in men – something that we can account for in our model too.

In our modelling we make a strong assumption that infection risks in population with comorbidities are the same as in the general population. In other words, we assume that denominator for IFR is same in both populations. We then specify a generic partial pooling model of fatality risks (FR, defined as number of deaths in the entire population, without regards to infection status) such that

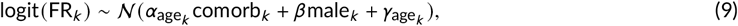

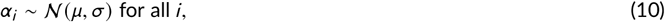

where, for *k* -th observation, age_*k*_ is the age group, and comorb_*k*_ and male_*k*_ are indicator variables. This means that each age group is assigned different “baseline” fatality risks.

Note that the assumption of FRs varying across age groups that we just mentioned differs from the model of age-specific IFRs in Section 2. This is because we hypothesised that infection risks (which are used as denominators in the IFR model of Section 2) will vary across age groups. This is borne out by Figure 5.

**FIGURE 5.**
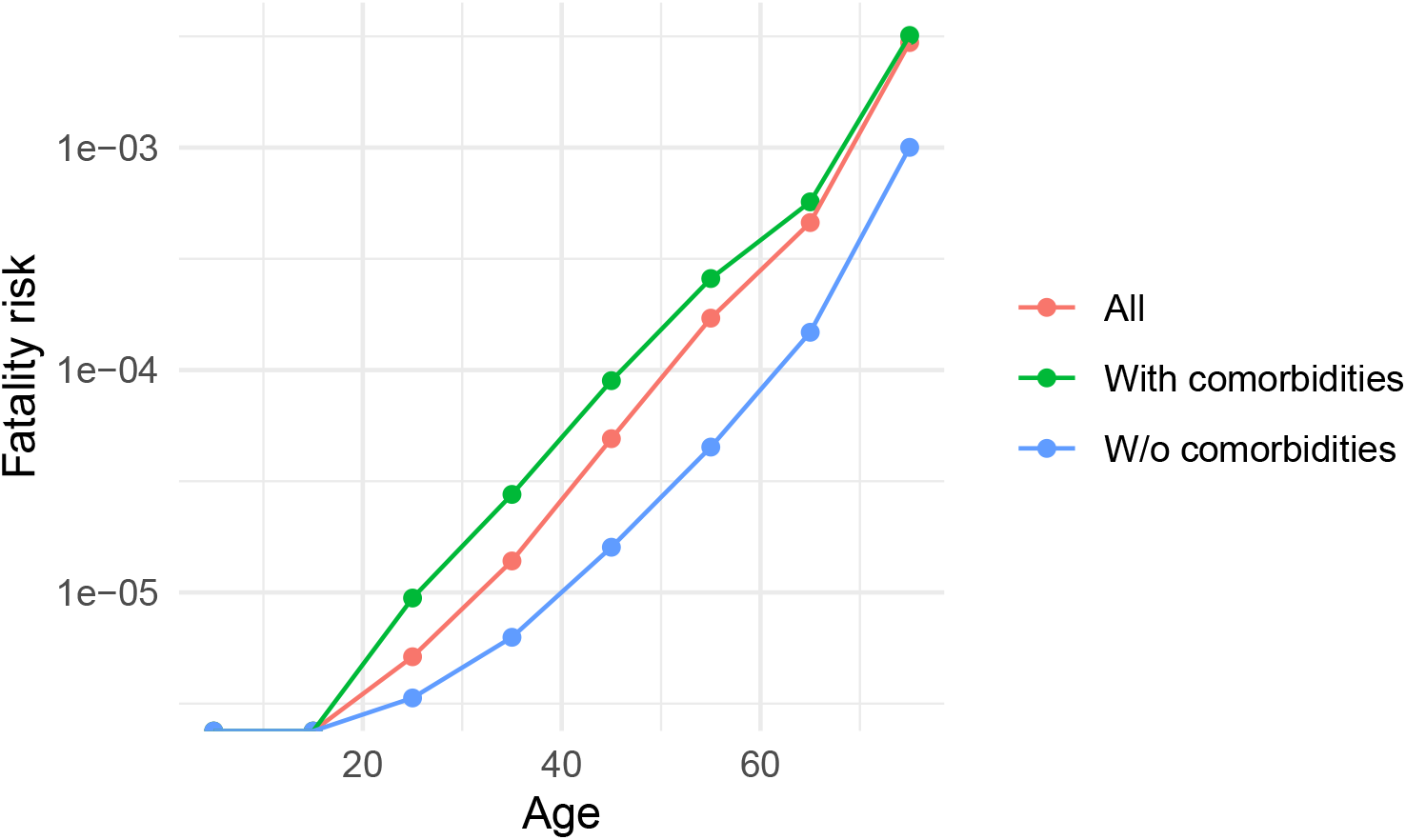
Fatality risks as a function of age in OpenSAFELY data. Fatality risk is zero in the 0-10 and 10-20 age groups.

**FIGURE 6.**
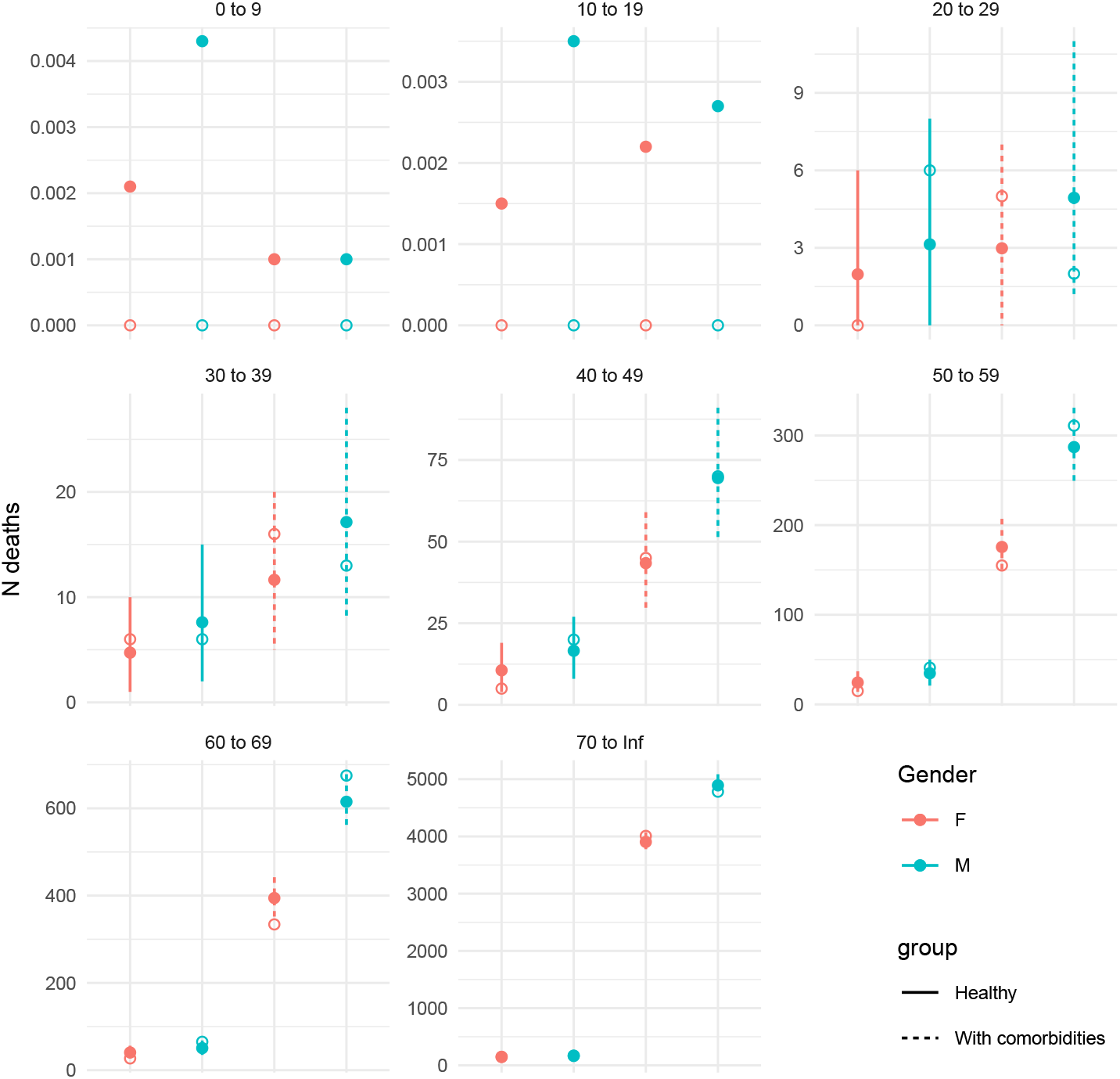
Comparison of posterior predictive numbers of deaths from the fitted Bayesian model (mean and 95% uncertainty intervals) with data inputs (circles). For each age grouping we have 4 estimates: male/female and healthy vs general population

**FIGURE 7.**
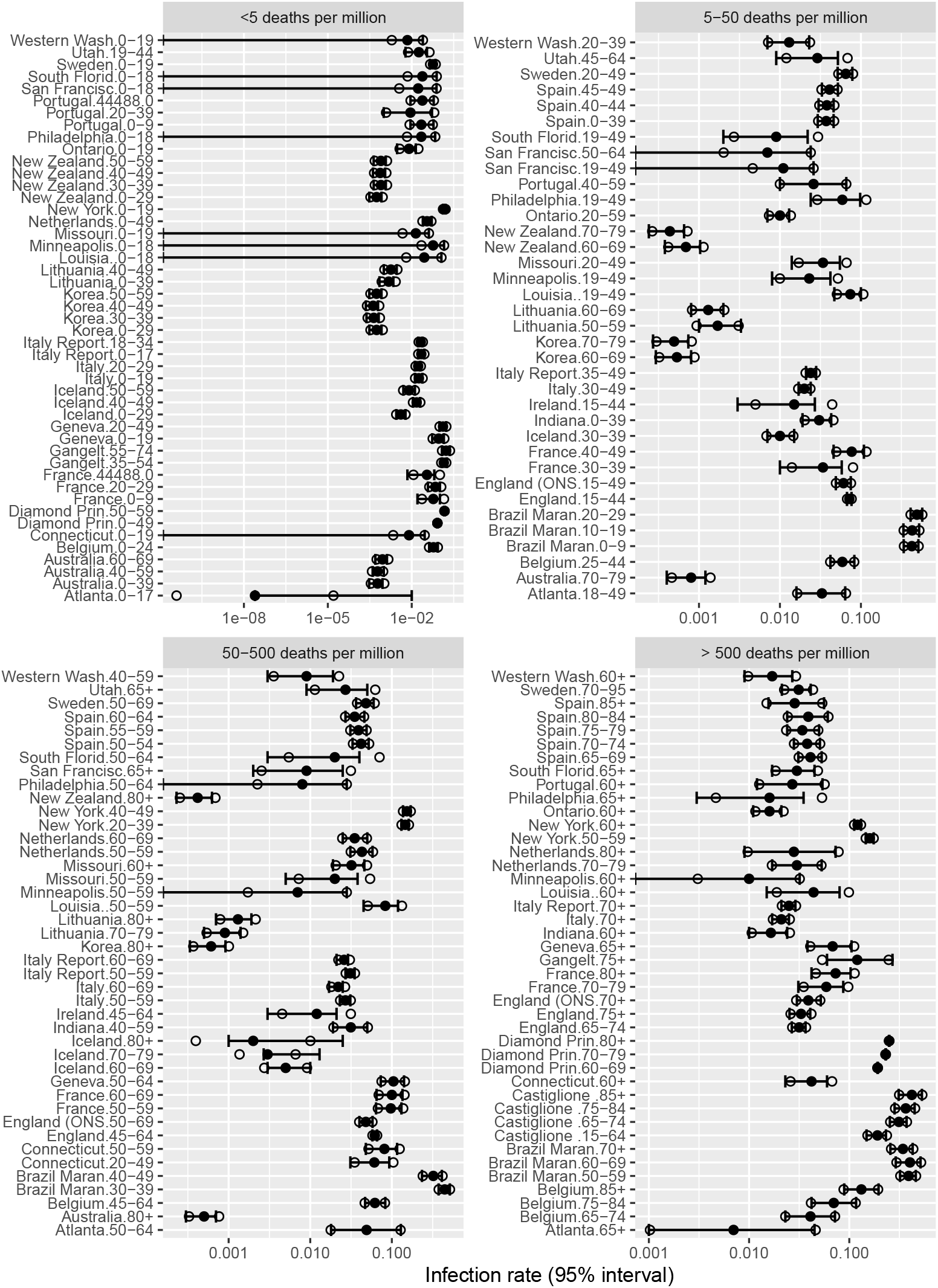
Comparison of model-estimated prevalences (95% CI’s reported by modelling studies) collected by Levin, Cochran, and Walsh (2020) and our distributional assumptions: additional circles show 95% CIs recreated by assuming logit-normal distribution of prevalence. We group studies into 4 bands of mortality to mirror earlier figures. Please note that this approach produces discrepancies in a number of US estimates where the confidence intervals were skewed toward including 0. However, since we do not have access to source data, we decided to use the logit-normal assumption for all estimates. This assumption may have an effect of overestimating mortality risk in settings where prevalence was very low.

Summary of the main model parameters is as follows:

~~~
##           mean    se_mean    sd    2.5%   25%   50%   75%   98%   n_eff Rhat
## mu        1.43    0.00278  0.190   1.05         1.33          1.43        1.53        1.81        4674         1
## sigma     0.36    0.00462  0.212   0.12         0.22          0.31        0.44        0.90        2115         1
## alpha[3]  1.33    0.00440  0.329   0.60         1.14          1.34        1.53        1.95        5577         1
## beta      0.40    0.00019  0.019   0.36         0.39          0.40        0.42        0.44        10314        1
## gamma[3]  −13.05  0.00415  0.342   −13.76       −13.27        −13.04       −12.82     −12.43      6795         1
~~~

We find that the mean risk ratio between population with comorbidities and healthy sub-population in 20-29 age group (exponent of *α*_3_ above) is 3.97, with wide 95% uncertainty interval of 1.81 to 7.05.^A8^

Next, using the posterior samples we calculate the event rate in total population (i.e. *θ*\* = (*θ*_1_*n*_1_ + *θ*_2_*n*_2_)/(*n*_1_ + *n*_2_), where subscripts 1 and 2 are healthy and comorbid sub-populations) and then divide it by ratio in healthy population to obtain an estimate of risk reduction possible by selecting healthy volunteers only. The mean posterior value is 1.88, with 95% uncertainty interval from 1.24 to 2.79. Due to use of Bayesian hierarchical model over many age groups, the uncertainty interval is much narrower than on the risk reduction factor calculated on 20-29 age group only.

To validate the model, we conducted a simple posterior predictive check for numbers of deaths in different age groups and genders. The graphical check is presented in Figure 6. Overall we find that the simple model has no problem with reproducing observed data.

### A-4 CONCLUSION AND SUMMARY OF RESULTS

In conclusion, the implications of the model for the risk in healthy young subjects are as follows:

- We find that average IFR in 20-29 age group for the studies included in this analysis is 1.51 × 10^−4^ with 95% interval from 1.18 × 10^−4^ to 1.92 × 10^−4^.
  – It is feasible that the mean IFR can be decreased as much as 3.82-fold (2*σ* impact on the IFR according to hyper-SD parameter in the meta-analysis model).
  – It is easy to argue that a HCT designer would be able to achieve IFR at least as low as within any of the large-scale studies included in our sample of populations. The smallest posterior mean for 20-29 year olds is 4.67 × 10^−5^, fitted to data from France.
  – Extending the HCT population to also include 30-39 year olds would lead to mean IFR of 2.65 × 10^−4^ with 95% interval from 2.06 × 10^−4^ to 3.35 × 10^−4^. Lowest mean IFR would then be 8.2 × 10^−5^ (also in France).
- In the general population the risk rises by the factor or 3.06 per each decade of age, with 95% interval from 3.01 to 3.11.
- In healthy population (defined as lack of co-morbidities listed above), the average mortality risk in 20-29 year olds is 1.88 times lower than in the general population, with 95% uncertainty interval from 1.24 to 2.79.
  – Our 1.88 estimate is a bit higher than the mean “crude” risk ratio of 1.53 because we use a Bayesian hierarchical model that synthesises evidence across all age groups.
  – Expanding to 20-39 year olds, the risk in healthy sub-population would be 2.04 times lower than in the general population (95% interval from 1.24 to 2.79).
- Combining the smallest posterior IFR for 20-29 year olds with our estimated fold-reduction due to excluding individuals with co-morbidities from the population would lead to a mean infection fatality risk of 2.6 × 10^−5^ with 95% Bayesian interval from 1.62 × 10^−5^ to 3.89 × 10^−5^.
  – In 20-39 year old subjects the risk would be 4.09 × 10^−5^ (2.88 × 10^−5^ to 5.55 × 10^−5^).

### A-5 SUMMARY OF INCLUDED STUDIES AND COMPLETE INPUT DATA

We present two tables, one listing all study-level information and another breaking down information by age group. References to all included studies are given in a separate bibliography at the end of this appendix. Below the table, we provide a list that can be used to cross-reference study locations with their bibliographic references.

Atlanta: Biggs et al. (2020); Australia: A. D. of Health (2020); Belgium: Herzog et al. (2020); Belgium: Molenberghs et al. (2020); Brazil Maranhao and Sao Luis: Silva et al. (2020); Brazil Regional: Hallal et al. (2020); Cache County, UT: Project (2020); Castiglione d’Adda: Pagani et al. (2020); Connecticut: Havers et al. (2020); Connecticut: Mahajan et al. (2020); Diamond Princess: Mizumoto et al. (2020); England: Ward et al. (2020); England (ONS): England (2020); France: Carrat et al. (2020); France: F. P. Health (2020); Gangelt: Streeck et al. (2020); Geneva: Perez-Saez et al. (2020); Iceland: I. D. of Health (2020); Indiana: Menachemi et al. (2020); Ireland: HPSC (2020); Italy: Istat (2020); Italy Deaths: Group (2020); Italy Report: Istat (2020); Korea: Control and Agency (2020); Lithuania: Registry (2020); Lombardy: Paradisi and Rinaldi (2020); Louisia.: Havers et al. (2020); Minneapolis: Havers et al. (2020); Missouri: Havers et al. (2020); Netherlands: RIVM (2020); New York: Rosenberg et al. (2020); New York State Comorbidity: N. Y. S. D. of Health (2020); New Zealand: N. Z. M. of Health (2020); NYC JAMA: Richardson S (2020); Ontario: Health Protection and Health Ontario) (2020); Philadelphia: Havers et al. (2020); Portugal: Saúde (2020); San Francisco Bay: Havers et al. (2020); South Florida: Havers et al. (2020); Spain: Pastor-Barriuso et al. (2020); Sweden: Authority (2020); Utah: Havers et al. (2020); Utah: Project (2020); Verity et al.: Verity, Okell, et al. (2020b); Washington County, UT: Project (2020); Weber County, UT: Project (2020); West Salt Lake, UT: Project (2020); Western Washington: Havers et al. (2020)

## Footnotes

1 Since this is only an assumption, rather than a fact which we can derive directly from data, we will present both the overall estimate across studies, or the expected lower risk. Similarly, users of our tool can choose between different assumptions.

2 Briefly, let *k* index populations and loc_*k*_ their locations (countries, regions). Let *d*_*k*_ be observed deaths for data point 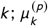 the reported mean prevalence and 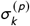 its standard error, on logit scale; *n*_*k*_ the total population; *X* is a vector of median ages, expressed in decades and centered at 25 years. The hierarchical model is as follows: 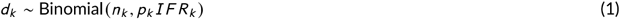 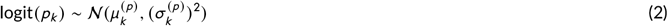 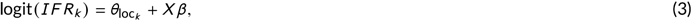 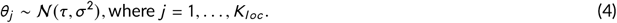 The parameters of the model are *p*_*k*_, the true prevalence; *θ*, location-specific (random) effects on IFR; *β*, (fixed) effect of age; *τ* and *σ*, the hyper-mean and hyper-scale parameters for IFR. We implement our model in Stan, (Carpenter et al., 2017) with weakly informative priors on all parameters. The prior for *τ* is centered at 1 death per 10,000 cases.

3 The lower-risk group is defined as non-obese, non-smoking, and without the following risk factors (following those used by (Williamson et al., 2020)): asthma, other chronic respiratory disease, chronic heart disease, diabetes mellitus, chronic liver disease, chronic neurological diseases, common autoimmune diseases (rheumatoid arthritis, systemic lupus erythematosus or psoriasis), solid organ transplant, asplenia, other immunosuppressive conditions, cancer, evidence of reduced kidney function, and raised blood pressure or a diagnosis of hypertension.

4 There is an assumption implicit in the model that the trial uses dose escalation or other data to ensure that the dose given does not greatly exceed the typical natural dose, or that a larger-than natural dose does not increase disease severity.

5 The assumed independence is conditional on the age and health status of participants, and for dose-response studies, also infection severity by dose.

6 This is equivalent to assuming that the risk of infection at each dose is above the threshold for infection to replace a dose-response curve.

7 See discussion below about incorporating other risks in the model as more data becomes available.

8 All intervals reported here are Bayesian posterior intervals. For brevity we just refer to them as “x% interval” or “uncertainty interval,” (Gelman, 2010).

9 Denoting by *σ* the hyper-scale parameter in the hierarchical model, 2*σ* impact corresponds to 3.96-fold mean decrease in IFR. That means we expect 2.5% of studies to have IFR more than 4 times lower than the average IFR.

A1 The role of time may be due to new treatments, improvements over time in our ability to treat COVID-19 or selection pressures which may lead to more benign versions of the virus. Country-specific or location-specific factors in IFR data may be driven by under-reporting, health care factors (including access to health care services) or underlying distributions of known risk factors. Additionally, some unknown risk factors (e.g. genetic) may also be operating, in which case controlling for age and co-morbidities will be not sufficient to account for cross-location differences.

A2 Various estimates published since suggested that the relationship of mortality risk to age is consistent across different countries.

A2 Various estimates published since suggested that the relationship of mortality risk to age is consistent across different countries.

A3 It is also possible to work with *I F R*_*k*_ parameters and treat them as derived from Beta distribution with some “hyperparameters” *α* and *β* Of Beta distribution, as done by e.g. Carpenter (2016). That approach, however, does not offer an easy way of modelling impact of covariates (e.g. age and co-morbidities) on the rates.

A4 Another advantage of such a model is that it can use either individual-level or summary data and work with covariates (such as gender, age, time of the study, co-morbidities), captured as odds ratios or risk ratios. If only summary data are available, covariates can be defined as study level distributions (e.g. % male)

A5 This basic approach potentially exaggerates uncertainty, as we treat different 95% intervals reported in the study as uncorrelated.

A6 “asthma, other chronic respiratory disease, chronic heart disease, diabetes mellitus, chronic liver disease, chronic neurological diseases, common autoimmune diseases (Rheumatoid Arthritis (RA), Systemic Lupus Erythematosus (SLE) or psoriasis), solid organ transplant, asplenia, other immunosuppressive conditions, cancer, evidence of reduced kidney function, and raised blood pressure or a diagnosis of hypertension”

A7 We obtain the interval using a simulation approach. Using normal approximation of log(RR) statistic we obtain a narrower 0.57 to 4.1, perhaps due to poor quality of approximation for rare events.

A8 Using simple models that assumed identical risk ratios in all age groups would lead to a mean RR of similar magnitude but a much less uncertain estimate, due to more rigid model assumptions; similarly, assuming some linear age structure on risks, such as in the main meta-analysis model above, would may lead to a different RR, but we do not think such an assumption is justified here. We do not include outputs of these models in this short write-up.

